# Beyond Rurality: Individual Socioeconomic Status and Chronic Disease Prevalence

**DOI:** 10.64898/2026.04.02.26350063

**Authors:** Shivani Sabarish, Chung-Il Wi, Madison Beenken, Dave Watson, Christi A. Patten, Tabetha A. Brockman, Christine M. Prissel, Philip Wheeler, Dan Kelleher, Gokhan Anil, Trent Anderson, Eunice Y. Park, Gurpreet Singh, Nahyr Lugo-Fagundo, James F. Howick V, Cheryl L. Walker-Mcgill, Brandon H. Hidaka, Pravesh Sharma, Sagar B. Dugani, Thanai Pongdee, Jessica L. Sosso, Randy M. Foss, Prathibha Varkey, Vesna D. Garovic, Young J. Juhn

## Abstract

**Importance:** Rural-urban disparities in chronic disease prevalence are well established; however, the extent to which individual-level socioeconomic status (SES) contributes to these disparities remains unclear.

**Objective:** To examine the associations of rurality and SES with the prevalence of five most burdensome chronic diseases among adults.

**Design:** We conducted a retrospective cross-sectional study of adults across 27 Upper Midwest counties using the Expanded Rochester Epidemiology Project (E-REP) medical record data linkage system to evaluate associations between rurality, SES and chronic disease prevalence. Prevalence of clinically diagnosed asthma, diabetes, hypertension, coronary heart disease, and mood disorders was identified from International Classification of Diseases ICD9/10 codes over a five-year period (2014-2019).

**Setting:** Population based

**Participants:** Adults over 18 years residing in the 27 E-REP counties, excluding those missing rural-urban residence status.

**Exposure:** HOUSES index, an individual-level measure of SES, served as the primary measure, while rurality based on Rural Urban Commuting Area (RUCA) codes 4-10 was the secondary measure.

**Main Outcome:** Prevalence of the five clinically diagnosed chronic diseases was identified using ICD9/10 codes from 2014–2019. Mixed effect logistic regression models were used and adjusted for demographics and general medical examination receipt, to assess rural-urban and SES differences for prevalence of each chronic disease.

**Results:** Among 455,802 adults with available HOUSES index, 42.8% lived in rural areas, 53.8% were female and 87.4% were non-Hispanic White. In the unadjusted analysis, rural and urban populations showed comparable asthma and CHD prevalence, while mood disorders, hypertension, and diabetes were more common in urban areas. After adjusting for demographic factors and healthcare utilization, rural-urban differences were no longer statistically significant, whereas SES remained strongly associated with all diseases in a dose response manner (e.g., adjusted Odds Ratio for hypertension (ref: HOUSES index Q4): 1.14, 1.27, and 1.42 for HOUSES index Q3, Q2, and Q1, respectively).

**Conclusions and Relevance:** Individual-level SES measured by the HOUSES index, was more strongly associated with chronic disease prevalence than rurality, supporting its integration into population health assessment and risk stratification.

## INTRODUCTION

Rurality is a complex, multidimensional, and inconsistently defined construct intended to capture geographic context, population density, economic structure, and access to services, yet it has remained a central focus of health disparities research largely because more precise measures of structural vulnerability have been difficult to operationalize.^1–3^ Existing rurality definitions vary widely across federal^4–6^ and research frameworks, limiting comparisons across studies.^7, 8^ For example, an individual may be categorized as living in a rural area based on Rural Urban Commuting Area (RUCA) codes but as urban using U.S. Census criteria, reflecting fundamental differences in the underlying classification logic.^9^ Moreover, rurality classifications mask substantial variations in socioeconomic conditions such as income, education, and wealth.^6, 10^ Although numerous studies have documented rural–urban differences in health outcomes,^11^ including higher chronic disease prevalence,^12^ and lower uptake of preventive services,^13^ other research demonstrates condition-specific or inconsistent associations between rurality and disease risk.^14^These discrepancies indicate that the rural–urban binary with different thresholds may oversimplify a construct that more accurately exists along a continuum rather than as a discrete category.^15, 16^

Socioeconomic status (SES) consistently emerges in literature as a major determinant of health outcomes and may exert a more substantial influence on disease patterns than rural residence alone.^17^ It is a more sensitive indicator of vulnerability because socioeconomic disadvantage is not geographically fixed^18^; urban areas may contain concentrated pockets of poverty,^19^ while some rural areas are comparatively affluent.^20^ Taken together, these findings emphasize the necessity of replacing broad rural–urban distinctions with a deeper analysis of socioeconomic stratification, including the specific SES differences present in both rural and urban settings.

Selecting an appropriate SES measure for research presents an ongoing challenge.^21^ SES can be operationalized in multiple ways; including individual-level indicators (e.g., income, education, occupation, insurance status, wealth)^22^ or contextual measures applied at broader geographic scales, (e.g., neighborhood-level, census tract-level, county-level).^23, 24^ However, some studies contend that relying on a univariate SES indicator may fail to capture the multidimensional nature of socioeconomic position.^21^ While composite measures such as the Area Deprivation Index (ADI) capture multiple socioeconomic dimensions,^25^ studies also note that these indices may not consistently reflect individual level social risk, underscoring persistent limitations in SES measurement.^26, 27^ To address these issues, we use the HOUSES index, a housing-based, individual-level SES measure that offers an alternative to neighborhood-based indices like ADI.^28, 29^ Whereas ADI assigns a uniform score to all residents of a given census block group, HOUSES captures SES at the level of the individual through property-level housing data.^29^

Although a substantial body of work has examined the association between individual SES indicators and chronic disease, far fewer investigate how these patterns vary across rural and urban settings.^30^ In contrast, rural–urban health research often treats SES merely as a covariate, commonly relying on single variable measures that inadequately represent socioeconomic complexity.^21^ In this paper, we aim to fill this gap by closely investigating SES differences across geographic contexts and their association with chronic disease.

This study aims to assess the extent to which individual SES, measured using the HOUSES index, accounts for rural-urban differences in chronic disease prevalence. By incorporating this measure, we aimed to more precisely characterize socioeconomic influences on health and to assess whether disparities typically attributed to rurality are better explained by underlying SES differences rather than geographic classification alone.

## Materials and Methods

### Study setting and population

The Expanded Rochester Epidemiology Project (E-REP) links medical records from major health systems serving a defined 27 county area spanning 19 counties in southern Minnesota and eight counties in western Wisconsin which includes Mayo Clinic, Mayo Clinic Health System, Olmsted Medical Center, and Olmsted County Public Health Services.^31^ Although the overall E-REP capture rate is approximately 61% of the 27 county population, coverage varies widely across the region from nearly complete capture in Olmsted County, MN (~99.9%) to as low as ~21% in Brown County, MN, because inclusion requires receiving care at a participating institution and, in Minnesota counties, providing research authorization. Not all healthcare facilities in the region participate in the E-REP, contributing to this geographic variation in population capture.^31^ The study population was drawn from this geographically defined region, which includes a mixture of rural and urban communities. The captured E-REP population was predominantly White (87.6%) and consisted of 51.4% females.^32, 33^

### Study Design

We conducted a population-based, retrospective cross-sectional study that evaluated the five-year prevalence (April 1, 2014, to April 1, 2019) of five common chronic diseases with significant health burden.

### Definition of Rurality

The term “rural” has a broad range of meanings, including agricultural regions, small towns, geographic isolation, and low population density,^34^ and different federal agencies use various definitions.^35^ For this study, we used the RUCA classification system, which was developed in the 1990s to provide a subcounty-level measure of rurality based on population density, urbanization, and commuting patterns. It assigns 10 primary codes reflecting metropolitan, micropolitan, small town, and rural areas, with additional secondary codes offering further granularity. ^36^ Federal guidelines define rurality using primary codes 4-10.^6^ Given that primary RUCA codes remain the standard practice^7, 37^ used in rural-urban health research and offer broad, interpretable categories, our analysis relied solely on these primary codes. Rurality was determined for each individual based on address in E-REP as of April 1, 2019.

### Socioeconomic status measured by HOUSES index

The HOUSES index is a tool developed to address limitations of area-level or neighborhood-level socioeconomic measures derived from census data, HOUSES is an individual-level SES measure created by investigators at Mayo Clinic and is based on objectively measured housing characteristics obtained from public property assessment records.^28, 29^ Residential addresses are geocoded and linked to county tax assessor data to extract four housing variables: assessed property value, square footage, number of bedrooms, and number of bathrooms. Each component is standardized to a z-score, and the composite HOUSES index is calculated as the sum of these standardized values. Scores are then categorized into quartiles, with Q1 representing the lowest SES and Q4 the highest. The HOUSES index has been validated across multiple studies as a reliable proxy for SES, particularly in settings where traditional SES metrics are unavailable, incomplete, or inaccurate. Prior research has demonstrated that lower HOUSES index are associated with a range of adverse health outcomes,^38^ including lower vaccination rates in children (e.g., pneumococcal,^39^ HPV^40^), higher likelihood of smoking in adolescents,^41^ and greater risk of falls,^42^ poorer depression treatment outcomes,^43^ and reduced diabetes control in adults.^44^ HOUSES index was determined for each individual as of April 1, 2019.

### Development of HIPAA-compliant, cloud-based HOUSES Platform

With support from a recently completed NIH-funded study (CT number: R21HD 51902-2), the HOUSES Platform was developed to make HOUSES index nationally scalable and HIPAA-compliant. The platform provides geocoded coordinates, HOUSES index, ADI, RUCA-based rural classifications, and distance to reference points (e.g., emergency rooms) via a secure, API-enabled cloud infrastructure.

### Prevalence of five chronic diseases

We estimated the five-year prevalence of diabetes mellitus, hypertension, mood disorders, asthma, and coronary heart disease (CHD) among adults aged ≥18 years using EHR data from April 1, 2014, to April 1, 2019. These five diseases were selected based on the Agency for Healthcare Research and Quality’s (AHRQ) designation of them as among the most burdensome chronic diseases.^14, 45^ Cases were ascertained using the E-REP, requiring at least one disease-specific International Classification of Disease-Clinical Modification (ICD-9-CM or ICD-10-CM) documented at any healthcare institution within the 27 included counties *(see Supplementary Material).* All ICD codes were selected based on clinical classification codes proposed by the AHRQ Healthcare Cost and Utilization Project.^45^ ICD codes are widely used in administrative and Electronic Health Record (EHR) based research because validation studies show that ICD derived chronic disease estimates typically differ from chart review by only 1-7%, supporting their continued use despite fluctuations in coding accuracy over time.^46^

### Other Variables

Demographics population characteristics included age (as of April 1, 2014), sex, and race and ethnicity. Race/ethnicity was recorded as non-Hispanic White, Black or African American, Hispanic or Latino, Asian, or other/ unknown, based on self-reported information in the EHR. Receipt of at least one general medical examination (GME) during the five-year period, identified using relevant Current Procedural Terminology codes, served as a surrogate marker of healthcare utilization.

### Statistical Analysis

Patient characteristics and distribution of disease prevalence were summarized using mean and standard deviation for continuous metrics and counts and percentages for categorical metrics. Differences between rural and urban patients were made using two-sample t-tests, chi-square tests, and corresponding p-values. Mixed effects logistic regression models with county as a random effect were used to examine the univariate association between rurality and the prevalence of each of the five diseases. Next, multivariable logistic models adjusted for demographic factors, including age, sex, and race/ethnicity, GME and known risk factors for diabetes (hyperlipidemia and obesity) and CHD (diabetes, hyperlipidemia, and obesity). Then we repeated the analyses with HOUSES as the main exposure, instead of rurality. Finally, we built models including both rurality and HOUSES to assess their associations with disease prevalence. Individuals with race/ethnicity recorded as “unknown” (<2%) were grouped into the “other” category for analysis. Results are reported as unadjusted and adjusted odds ratios with corresponding 95% confidence intervals. Statistical significance was defined as p < 0.05. All analyses were conducted using R (R Core Team, 2024).

## Results

### Basic characteristics of subjects

A total of 476,767 patients were identified in the 27 county E-REP area from 2014 to 2019 and 20,965 (4.4%) were excluded due to missing or invalid RUCA codes. Rural residents were older than urban residents, on average (52.2 vs 49.5 years). The rural population had a higher proportion of White individuals (88.9% vs 86.4%) and lower proportions of Asian (1.0% vs 3.0%) and Black (1.7% vs 3.5%) individuals, while the proportion of individuals identifying as Hispanic was slightly higher in rural areas (5.5% vs 4.1%). SES also differed by rurality, with rural residents more frequently represented in the lower quartile (Q1: 21.7% vs 19.8%) and less represented in the highest quartile (Q4: 26.6% vs 32.1%). Health care utilization was lower among rural residents, with fewer having at least one GME in the five-year period (55.5% vs 57.0%). (Table 1)

**Table 1.**
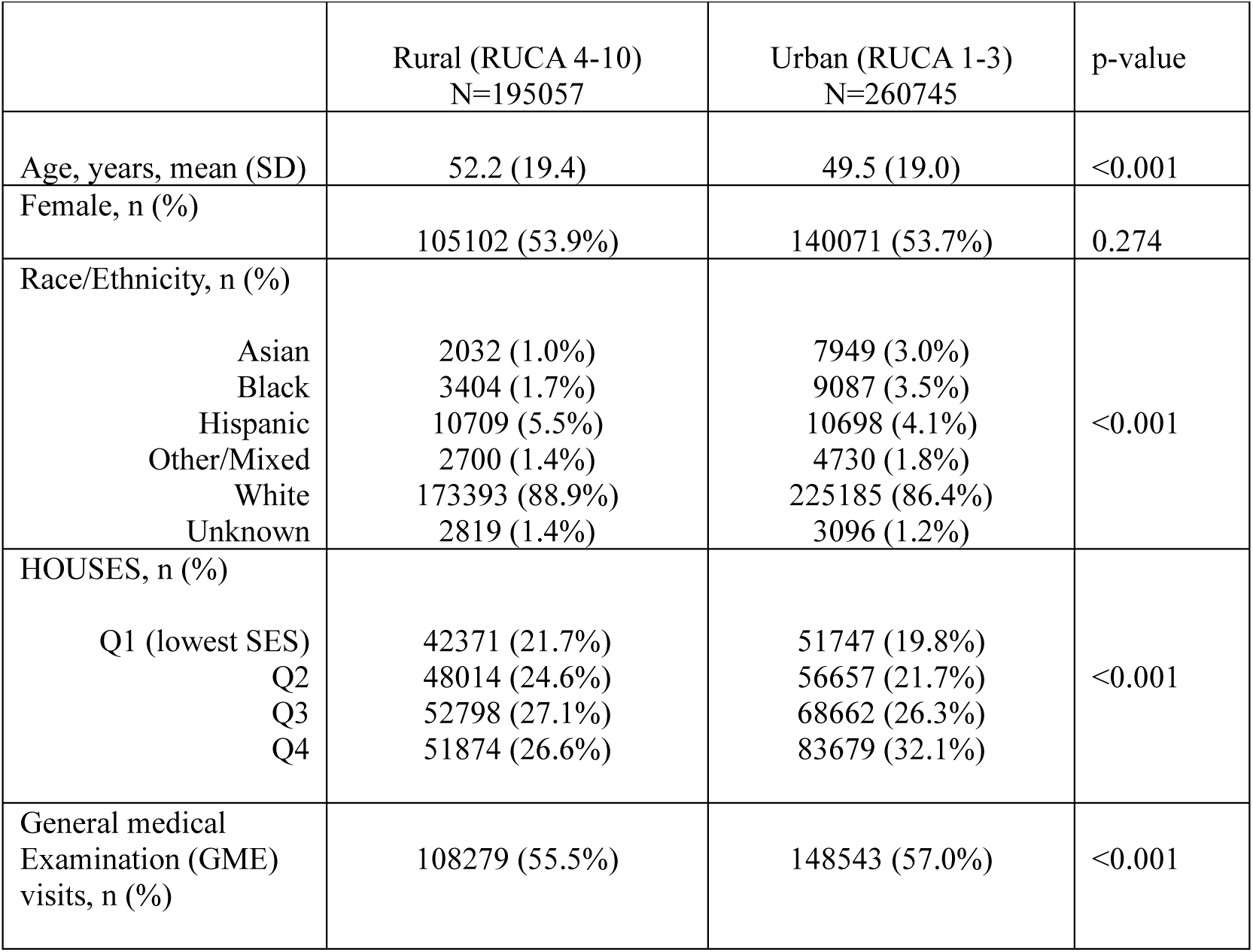
Basic characteristics of study subjects by rurality (RUCA)

### Rurality and Prevalence of Chronic diseases without adjusting for SES

Table 2 shows the crude prevalence of the five chronic diseases by rurality, which showed residents of rural areas exhibited higher prevalence for all diseases except asthma, which was more common among urban residents. Consistent with these crude patterns, the unadjusted regression model (Model 0) indicated that rural residence was associated with higher odds of all chronic diseases except asthma.

**Table 2:** Crude Prevalence Estimates and Odds Ratios^a^ Stratified by Rural and Urban Residence.

|  |  |  |  |
| --- | --- | --- | --- |
| **Prevalence** | **Measure: Rurality** | **RUCA** |  |
|  |  | **Prevalence (%), p value** | **Model 0<sup>a</sup>** |
| **Asthma** | Rural<br>Urban | 16581 (8.5%), 0.030<br>22639 (8.7%) | Ref<br>0.99 (0.95, 1.02) |
| **Mood Disorder** | Rural<br>Urban | 60871 (31.2%), <0.001<br>78121 (30.0%) | Ref<br>0.97 (0.94, 0.99) |
| **Hypertension** | Rural<br>Urban | 22027 (11.3%), <0.001<br>22675 (8.7%) | Ref<br>0.91 (0.89, 0.93) |
| **Diabetes** | Rural<br>Urban | 29935 (15.3%), <0.001<br>35383 (13.6%) | Ref<br>0.87 (0.85, 0.90) |
| **CHD** | Rural<br>Urban | 28664 (14.7%), <0.001<br>33253 (12.8%) | Ref<br>0.93 (0.90, 0.96) |
| RUCA: Rural Urban Commuting Area |  |  |  |
| <sup>a</sup>Model 0: Univariate analysis for prevalence (unadjusted OR) (independent variable: RUCA) |  |  |  |

In adjusted analyses, as shown in Table 3, (Model 1; controlling for age, sex, and GME), urban residence was associated with lower odds of mood disorders (OR = 0.95, 95% CI: 0.92–0.97) and lower odds of diabetes (0.92, 0.89–0.95). No significant adjusted associations were observed for asthma, hypertension, or CHD.

**Table 3:**
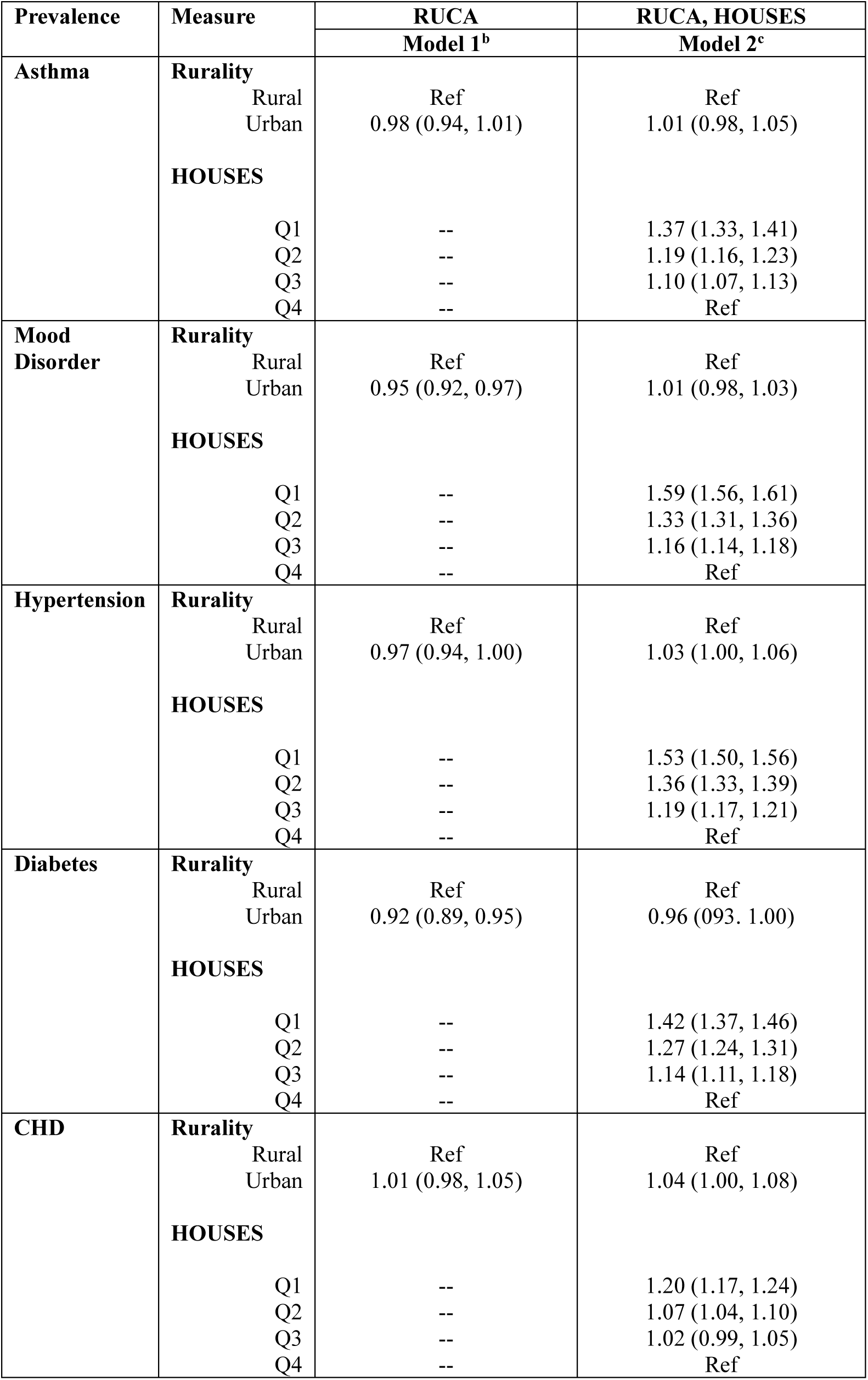
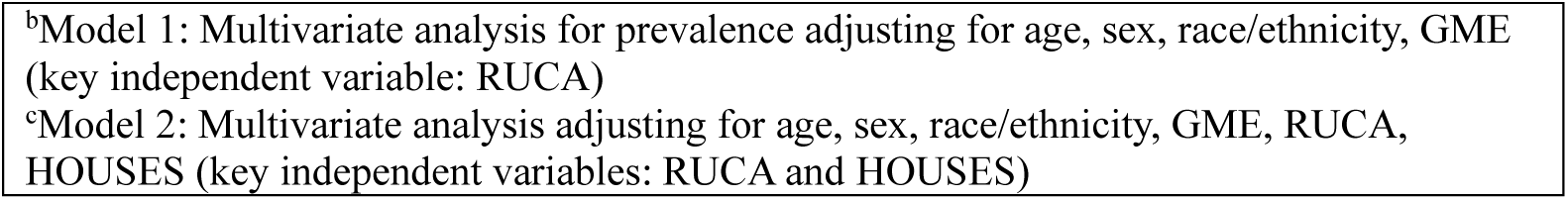
Association between Socioeconomic Status and Urban-Rural Status in Disease Prevalence.

### Rurality and Prevalence of Chronic diseases with adjusting for SES

In Table 3, we examined the extent to which SES explains rural-urban differences in disease prevalence. After adjustment for RUCA, HOUSES index, and all covariates, the associations between RUCA and disease prevalence were substantially attenuated or no longer significant. In contrast, HOUSES index remained strongly associated with every disease. Individuals in the lowest quartile (Q1) had higher odds compared with Q4 for asthma (1.37, 1.33-1.41), mood disorders (1.59, 1.56-1.61), hypertension (1.53, 1.50-1.56), diabetes (1.42, 1.37-1.46), and CHD (1.20, 1.17-1.24). These results indicate that SES largely account for rural–urban disparities.

For each disease, the interaction between rural status and SES was tested to assess differences in rural associations across SES quartiles. The interaction tests for mood disorders and asthma were statistically significant. *(See Supplementary material for rural versus urban odds ratios within SES quartiles)*

## DISCUSSION

In this population-based cross-sectional study of adults in the U.S. Midwest, we examined associations between rurality, individual-level SES, and the prevalence of five chronic diseases. In initial models that adjusted for demographic factors and healthcare utilization, urban residence was associated with modestly lower odds of mood disorder (OR = 0.95, 95% CI: 0.92-0.97) and diabetes (0.92, 0.89-0.95). However, after adjustment for individual-level SES measured by the HOUSES index, rural-urban differences in prevalence across all five diseases were substantially attenuated and no longer statistically significant, whereas the HOUSES index showed a consistent, graded dose-response relationship with each disease independent of rurality. Our findings indicate that individual-level SES shows a stronger association with chronic disease prevalence than rurality, although causality cannot be inferred.

Asthma was the only disease that did not favor either rural or urban residents across models. This aligns with the broader literature that reports mixed findings across rural-urban comparison.^35^ Some studies report higher asthma prevalence in rural areas,^47, 48^ others in urban settings,^49^ and some note higher mortality in rural populations.^50^

Our results also align with international and U.S. evidence showing SES being a stronger and more consistent predictor of chronic disease outcomes than geographical classifications. In the Prospective Urban Rural Epidemiology (PURE) study spanning 20 countries, SES measured by education was shown to be a more powerful predictor of cardiovascular outcomes than place of residence.^51^ Similarly, a recent 41 state Behavioral Risk Factor Surveillance System (BRFSS) analysis reported that an apparent rural–urban disparity in diabetes prevalence (14.3% vs 11.2%) was largely attenuated to non-significance after adjustment for SES (education, income) and obesity.^30^ Complementing these patterns, a large Finnish multi-cohort study with UK replication linked lower SES (area deprivation, education, occupational grade) to increased risk for 18 of 56 conditions, with early and strong associations for mental/behavioral disorders that preceded a cascade of physical diseases.^52^ Finally, a retrospective study in Olmsted County using the HOUSES index, found that lower SES was consistently associated with higher prevalence of five diseases (asthma, diabetes, hypertension, CHD and mood disorders) although SES-ethnicity interactions varied by condition.^14^

These findings have important implications for research and clinical care. Incorporating individual-level socioeconomic measures such as the HOUSES index may enable more precise identification of socioeconomically vulnerable patients whose disease burden may otherwise be attributed to geography alone. Lower SES populations consistently demonstrate reduced healthcare engagement, including delayed or underdiagnosis of chronic diseases,^53^ and higher rates of being lost to follow-up.^54^ If integrated into electronic health record systems, HOUSES index could support healthcare professionals, care coordinators, community health workers, and patient navigators in proactively identifying patients at socioeconomic risk and facilitating earlier referral pathways,^55^ targeted screening efforts,^56^ and SES-informed risk stratification within chronic disease management.^57^

### Strengths and Limitations

Our study is one of the largest, population-based studies (~400,000 individuals) that provides strong statistical power and enhances generalizability. Reliance on electronic health records ensured physician verified diagnoses rather than self-reported conditions, reducing misclassification, recall bias, and reporting error while offering objective clinical detail. The HOUSES index served as an alternative individual-level SES measure, offering an individualized and scalable approach that helps address some limitations of commonly used indicators such as area-level metrics, income, or education. Additionally, the near complete geocoding of residents included in REP minimized missing SES data and improved analytic precision.

Several limitations warrant caution in interpreting the findings. An important source of potential bias is the exclusion of 20,965 patients (4.4%) with missing or invalid RUCA codes and those without valid residential addresses for HOUSES scoring. These individuals may systematically differ from the analytic sample. A supplementary sensitivity analysis comparing included vs. excluded patients is recommended. The cohort was predominantly White, which limits applicability of these results to more racially and ethnically diverse populations. Despite covering a broad 27 county area, E-REP captures only patients seen within participating health systems, leading to partial and geographically variable population representation. Variability in institutional participation and data collection practices may further affect data consistency. As a result, people with low care access or utilization may be underrepresented, which could bias estimates of disease burden and health service use.^32, 33^ The use of EHR data introduces known biases, including underrepresentation of vulnerable groups and informed presence bias,^58, 59^ lack of consensus on documentation,^60^ especially with the provider-centric setup with limited input from the patients.^61^ Our findings may be affected by the 2015 ICD-9-CM to ICD-10-CM transition, which can artificially distort prevalence trends due to mapping misalignment and site-specific coding variability.^62^ Additionally, we used GME visits as a proxy for healthcare access; however, the cross-sectional design means GME visits could occur before or after disease onset, introducing temporal ambiguity that limits interpretation of GME as a strictly pre-diagnosis access measure. We used RUCA codes to distinguish between rural and urban areas, which, despite offering more granularity than county-level measures, still reduce rurality to a dichotomy. Therefore, further research is needed that operationalizes rurality using multiple categories rather than a dichotomous variable. Finally, a major limitation is that the cross-sectional study design precludes causal inference and limits evaluation of temporal relationships; longitudinal studies in more diverse populations are needed to validate and extend these findings.

## Conclusion

Overall, our findings reinforce that individual-level SES, rather than rural-urban residence alone, is the dominant determinant of chronic disease disparities and should be central to future clinical, public health, and policy strategies aimed at improving population health.

## Data Availability

All data produced in the present work are contained in the manuscript

## Acknowledgements

We are grateful to Ms. Kelly Okeson for her careful proofreading and editorial support. We also acknowledge the use of Microsoft 365 Copilot (version: bizchat.20260303.10.1), which aided in improving writing quality and clarity during manuscript preparation. The authors take responsibility for the integrity of the content generated.

## Declarations

### Author Approval

All authors reviewed and approved the final manuscript prior to submission.

### Ethics approval

This study was reviewed and approved by the Institutional Review Board Mayo clinic. (Approval number:06-009617)

### Data availability

The data supporting the findings of this study are available from the corresponding author upon reasonable request.

### Preprint statement

This manuscript has not been previously published, is not under consideration for publication elsewhere, and has not been posted on another preprint server.

## SUPPLEMENTARY MATERIAL

### Internal classification of diseases 9 and 10

#### ICD-10-CM

##### Diabetes

|  |  |
| --- | --- |
| *E0800* | *Diabetes mellitus due to underlying condition with hyperosmolarity without nonketotic hyperglycemic-hyperosmolar coma (NKHHC)* |
| *E0801* | Diabetes mellitus due to underlying condition with hyperosmolarity with coma |
| *E0810* | Diabetes mellitus due to underlying condition with ketoacidosis without coma |
| *E0811* | Diabetes mellitus due to underlying condition with ketoacidosis with coma |
| *E0821* | Diabetes mellitus due to underlying condition with diabetic nephropathy |
| *E0822* | Diabetes mellitus due to underlying condition with diabetic chronic kidney disease |
| *E0829* | Diabetes mellitus due to underlying condition with other diabetic kidney complication |
| *E08311* | Diabetes mellitus due to underlying condition with unspecified diabetic retinopathy with macular edema |
| *E08319* | Diabetes mellitus due to underlying condition with unspecified diabetic retinopathy without macular edema |
| *E08321* | Diabetes mellitus due to underlying condition with mild nonproliferative diabetic retinopathy with macular edema |
| *E083211* | Diabetes mellitus due to underlying condition with mild nonproliferative diabetic retinopathy with macular edema, right eye |
| *E083212* | Diabetes mellitus due to underlying condition with mild nonproliferative diabetic retinopathy with macular edema, left eye |
| *E083213* | Diabetes mellitus due to underlying condition with mild nonproliferative diabetic retinopathy with macular edema, bilateral |
| *E083219* | Diabetes mellitus due to underlying condition with mild nonproliferative diabetic retinopathy with macular edema, unspecified eye |
| *E08329* | Diabetes mellitus due to underlying condition with mild nonproliferative diabetic retinopathy without macular edema |
| *E083291* | Diabetes mellitus due to underlying condition with mild nonproliferative diabetic retinopathy without macular edema, right eye |
| *E083292* | Diabetes mellitus due to underlying condition with mild nonproliferative diabetic retinopathy without macular edema, left eye |
| *E083293* | Diabetes mellitus due to underlying condition with mild nonproliferative diabetic retinopathy without macular edema, bilateral |
| *E083299* | Diabetes mellitus due to underlying condition with mild nonproliferative diabetic retinopathy without macular edema, unspecified eye |
| *E08331* | Diabetes mellitus due to underlying condition with moderate nonproliferative diabetic retinopathy with macular edema |
| *E083311* | Diabetes mellitus due to underlying condition with moderate nonproliferative diabetic retinopathy with macular edema, right eye |
| *E083312* | Diabetes mellitus due to underlying condition with moderate nonproliferative diabetic retinopathy with macular edema, left eye |
| *E083313* | Diabetes mellitus due to underlying condition with moderate nonproliferative diabetic retinopathy with macular edema, bilateral |
| *E083319* | Diabetes mellitus due to underlying condition with moderate nonproliferative diabetic retinopathy with macular edema, unspecified eye |
| *E08339* | Diabetes mellitus due to underlying condition with moderate nonproliferative diabetic retinopathy without macular edema |
| *E083391* | Diabetes mellitus due to underlying condition with moderate nonproliferative diabetic retinopathy without macular edema, right eye |
| *E083392* | Diabetes mellitus due to underlying condition with moderate nonproliferative diabetic retinopathy without macular edema, left eye |
| *E083393* | Diabetes mellitus due to underlying condition with moderate nonproliferative diabetic retinopathy without macular edema, bilateral |
| *E083399* | Diabetes mellitus due to underlying condition with moderate nonproliferative diabetic retinopathy without macular edema, unspecified eye |
| *E08341* | Diabetes mellitus due to underlying condition with severe nonproliferative diabetic retinopathy with macular edema |
| *E083411* | Diabetes mellitus due to underlying condition with severe nonproliferative diabetic retinopathy with macular edema, right eye |
| *E083412* | Diabetes mellitus due to underlying condition with severe nonproliferative diabetic retinopathy with macular edema, left eye |
| *E083413* | Diabetes mellitus due to underlying condition with severe nonproliferative diabetic retinopathy with macular edema, bilateral |
| *E083419* | Diabetes mellitus due to underlying condition with severe nonproliferative diabetic retinopathy with macular edema, unspecified eye |
| *E08349* | Diabetes mellitus due to underlying condition with severe nonproliferative diabetic retinopathy without macular edema |
| *E083491* | Diabetes mellitus due to underlying condition with severe nonproliferative diabetic retinopathy without macular edema, right eye |
| *E083492* | Diabetes mellitus due to underlying condition with severe nonproliferative diabetic retinopathy without macular edema, left eye |
| *E083493* | Diabetes mellitus due to underlying condition with severe nonproliferative diabetic retinopathy without macular edema, bilateral |
| *E083499* | Diabetes mellitus due to underlying condition with severe nonproliferative diabetic retinopathy without macular edema, unspecified eye |
| *E08351* | Diabetes mellitus due to underlying condition with proliferative diabetic retinopathy with macular edema |
| *E083511* | Diabetes mellitus due to underlying condition with proliferative diabetic retinopathy with macular edema, right eye |
| *E083512* | Diabetes mellitus due to underlying condition with proliferative diabetic retinopathy with macular edema, left eye |
| *E083513* | Diabetes mellitus due to underlying condition with proliferative diabetic retinopathy with macular edema, bilateral |
| *E083519* | Diabetes mellitus due to underlying condition with proliferative diabetic retinopathy with macular edema, unspecified eye |
| *E083521* | Diabetes mellitus due to underlying condition with proliferative diabetic retinopathy with traction retinal detachment involving the macula, right eye |
| *E083522* | Diabetes mellitus due to underlying condition with proliferative diabetic retinopathy with traction retinal detachment involving the macula, left eye |
| *E083523* | Diabetes mellitus due to underlying condition with proliferative diabetic retinopathy with traction retinal detachment involving the macula, bilateral |
| *E083529* | Diabetes mellitus due to underlying condition with proliferative diabetic retinopathy with traction retinal detachment involving the macula, unspecified eye |
| *E083531* | Diabetes mellitus due to underlying condition with proliferative diabetic retinopathy with traction retinal detachment not involving the macula, right eye |
| *E083532* | Diabetes mellitus due to underlying condition with proliferative diabetic retinopathy with traction retinal detachment not involving the macula, left eye |
| *E083533* | Diabetes mellitus due to underlying condition with proliferative diabetic retinopathy with traction retinal detachment not involving the macula, bilateral |
| *E083539* | Diabetes mellitus due to underlying condition with proliferative diabetic retinopathy with traction retinal detachment not involving the macula, unspecified eye |
| *E083541* | Diabetes mellitus due to underlying condition with proliferative diabetic retinopathy with combined traction retinal detachment and rhegmatogenous retinal detachment, right eye |
| *E083542* | Diabetes mellitus due to underlying condition with proliferative diabetic retinopathy with combined traction retinal detachment and rhegmatogenous retinal detachment, left eye |
| *E083543* | Diabetes mellitus due to underlying condition with proliferative diabetic retinopathy with combined traction retinal detachment and rhegmatogenous retinal detachment, bilateral |
| *E083549* | Diabetes mellitus due to underlying condition with proliferative diabetic retinopathy with combined traction retinal detachment and rhegmatogenous retinal detachment, unspecified eye |
| *E083551* | Diabetes mellitus due to underlying condition with stable proliferative diabetic retinopathy, right eye |
| *E083552* | Diabetes mellitus due to underlying condition with stable proliferative diabetic retinopathy, left eye |
| *E083553* | Diabetes mellitus due to underlying condition with stable proliferative diabetic retinopathy, bilateral |
| *E083559* | Diabetes mellitus due to underlying condition with stable proliferative diabetic retinopathy, unspecified eye |
| *E08359* | Diabetes mellitus due to underlying condition with proliferative diabetic retinopathy without macular edema |
| *E083591* | Diabetes mellitus due to underlying condition with proliferative diabetic retinopathy without macular edema, right eye |
| *E083592* | Diabetes mellitus due to underlying condition with proliferative diabetic retinopathy without macular edema, left eye |
| *E083593* | Diabetes mellitus due to underlying condition with proliferative diabetic retinopathy without macular edema, bilateral |
| *E083599* | Diabetes mellitus due to underlying condition with proliferative diabetic retinopathy without macular edema, unspecified eye |
| *E0836* | Diabetes mellitus due to underlying condition with diabetic cataract |
| *E0837X1* | Diabetes mellitus due to underlying condition with diabetic macular edema, resolved following treatment, right eye |
| *E0837X2* | Diabetes mellitus due to underlying condition with diabetic macular edema, resolved following treatment, left eye |
| *E0837X3* | Diabetes mellitus due to underlying condition with diabetic macular edema, resolved following treatment, bilateral |
| *E0837X9* | Diabetes mellitus due to underlying condition with diabetic macular edema, resolved following treatment, unspecified eye |
| *E0839* | Diabetes mellitus due to underlying condition with other diabetic ophthalmic complication |
| *E0840* | Diabetes mellitus due to underlying condition with diabetic neuropathy, unspecified |
| *E0841* | Diabetes mellitus due to underlying condition with diabetic mononeuropathy |
| *E0842* | Diabetes mellitus due to underlying condition with diabetic polyneuropathy |
| *E0843* | Diabetes mellitus due to underlying condition with diabetic autonomic (poly)neuropathy |
| *E0844* | Diabetes mellitus due to underlying condition with diabetic amyotrophy |
| *E0849* | Diabetes mellitus due to underlying condition with other diabetic neurological complication |
| *E0851* | Diabetes mellitus due to underlying condition with diabetic peripheral angiopathy without gangrene |
| *E0852* | Diabetes mellitus due to underlying condition with diabetic peripheral angiopathy with gangrene |
| *E0859* | Diabetes mellitus due to underlying condition with other circulatory complications |
| *E08610* | Diabetes mellitus due to underlying condition with diabetic neuropathic arthropathy |
| *E08618* | Diabetes mellitus due to underlying condition with other diabetic arthropathy |
| *E08620* | Diabetes mellitus due to underlying condition with diabetic dermatitis |
| *E08621* | Diabetes mellitus due to underlying condition with foot ulcer |
| *E08622* | Diabetes mellitus due to underlying condition with other skin ulcer |
| *E08628* | Diabetes mellitus due to underlying condition with other skin complications |
| *E08630* | Diabetes mellitus due to underlying condition with periodontal disease |
| *E08638* | Diabetes mellitus due to underlying condition with other oral complications |
| *E08641* | Diabetes mellitus due to underlying condition with hypoglycemia with coma |
| *E08649* | Diabetes mellitus due to underlying condition with hypoglycemia without coma |
| *E0865* | Diabetes mellitus due to underlying condition with hyperglycemia |
| *E0869* | Diabetes mellitus due to underlying condition with other specified complication |
| *E088* | Diabetes mellitus due to underlying condition with unspecified complications |
| *E0900* | Drug or chemical induced diabetes mellitus with hyperosmolarity without nonketotic hyperglycemic-hyperosmolar coma (NKHHC) |
| *E0901* | Drug or chemical induced diabetes mellitus with hyperosmolarity with coma |
| *E0910* | Drug or chemical induced diabetes mellitus with ketoacidosis without coma |
| *E0911* | Drug or chemical induced diabetes mellitus with ketoacidosis with coma |
| *E0921* | Drug or chemical induced diabetes mellitus with diabetic nephropathy |
| *E0922* | Drug or chemical induced diabetes mellitus with diabetic chronic kidney disease |
| *E0929* | Drug or chemical induced diabetes mellitus with other diabetic kidney complication |
| *E09311* | Drug or chemical induced diabetes mellitus with unspecified diabetic retinopathy with macular edema |
| *E09319* | Drug or chemical induced diabetes mellitus with unspecified diabetic retinopathy without macular edema |
| *E09321* | Drug or chemical induced diabetes mellitus with mild nonproliferative diabetic retinopathy with macular edema |
| *E093211* | Drug or chemical induced diabetes mellitus with mild nonproliferative diabetic retinopathy with macular edema, right eye |
| *E093212* | Drug or chemical induced diabetes mellitus with mild nonproliferative diabetic retinopathy with macular edema, left eye |
| *E093213* | Drug or chemical induced diabetes mellitus with mild nonproliferative diabetic retinopathy with macular edema, bilateral |
| *E093219* | Drug or chemical induced diabetes mellitus with mild nonproliferative diabetic retinopathy with macular edema, unspecified eye |
| *E09329* | Drug or chemical induced diabetes mellitus with mild nonproliferative diabetic retinopathy without macular edema |
| *E093291* | Drug or chemical induced diabetes mellitus with mild nonproliferative diabetic retinopathy without macular edema, right eye |
| *E093292* | Drug or chemical induced diabetes mellitus with mild nonproliferative diabetic retinopathy without macular edema, left eye |
| *E093293* | Drug or chemical induced diabetes mellitus with mild nonproliferative diabetic retinopathy without macular edema, bilateral |
| *E093299* | Drug or chemical induced diabetes mellitus with mild nonproliferative diabetic retinopathy without macular edema, unspecified eye |
| *E09331* | Drug or chemical induced diabetes mellitus with moderate nonproliferative diabetic retinopathy with macular edema |
| *E093311* | Drug or chemical induced diabetes mellitus with moderate nonproliferative diabetic retinopathy with macular edema, right eye |
| *E093312* | Drug or chemical induced diabetes mellitus with moderate nonproliferative diabetic retinopathy with macular edema, left eye |
| *E093313* | Drug or chemical induced diabetes mellitus with moderate nonproliferative diabetic retinopathy with macular edema, bilateral |
| *E093319* | Drug or chemical induced diabetes mellitus with moderate nonproliferative diabetic retinopathy with macular edema, unspecified eye |
| *E09339* | Drug or chemical induced diabetes mellitus with moderate nonproliferative diabetic retinopathy without macular edema |
| *E093391* | Drug or chemical induced diabetes mellitus with moderate nonproliferative diabetic retinopathy without macular edema, right eye |
| *E093392* | Drug or chemical induced diabetes mellitus with moderate nonproliferative diabetic retinopathy without macular edema, left eye |
| *E093393* | Drug or chemical induced diabetes mellitus with moderate nonproliferative diabetic retinopathy without macular edema, bilateral |
| *E093399* | Drug or chemical induced diabetes mellitus with moderate nonproliferative diabetic retinopathy without macular edema, unspecified eye |
| *E09341* | Drug or chemical induced diabetes mellitus with severe nonproliferative diabetic retinopathy with macular edema |
| *E093411* | Drug or chemical induced diabetes mellitus with severe nonproliferative diabetic retinopathy with macular edema, right eye |
| *E093412* | Drug or chemical induced diabetes mellitus with severe nonproliferative diabetic retinopathy with macular edema, left eye |
| *E093413* | Drug or chemical induced diabetes mellitus with severe nonproliferative diabetic retinopathy with macular edema, bilateral |
| *E093419* | Drug or chemical induced diabetes mellitus with severe nonproliferative diabetic retinopathy with macular edema, unspecified eye |
| *E09349* | Drug or chemical induced diabetes mellitus with severe nonproliferative diabetic retinopathy without macular edema |
| *E093491* | Drug or chemical induced diabetes mellitus with severe nonproliferative diabetic retinopathy without macular edema, right eye |
| *E093492* | Drug or chemical induced diabetes mellitus with severe nonproliferative diabetic retinopathy without macular edema, left eye |
| *E093493* | Drug or chemical induced diabetes mellitus with severe nonproliferative diabetic retinopathy without macular edema, bilateral |
| *E093499* | Drug or chemical induced diabetes mellitus with severe nonproliferative diabetic retinopathy without macular edema, unspecified eye |
| *E09351* | Drug or chemical induced diabetes mellitus with proliferative diabetic retinopathy with macular edema |
| *E093511* | Drug or chemical induced diabetes mellitus with proliferative diabetic retinopathy with macular edema, right eye |
| *E093512* | Drug or chemical induced diabetes mellitus with proliferative diabetic retinopathy with macular edema, left eye |
| *E093513* | Drug or chemical induced diabetes mellitus with proliferative diabetic retinopathy with macular edema, bilateral |
| *E093519* | Drug or chemical induced diabetes mellitus with proliferative diabetic retinopathy with macular edema, unspecified eye |
| *E093521* | Drug or chemical induced diabetes mellitus with proliferative diabetic retinopathy with traction retinal detachment involving the macula, right eye |
| *E093522* | Drug or chemical induced diabetes mellitus with proliferative diabetic retinopathy with traction retinal detachment involving the macula, left eye |
| *E093523* | Drug or chemical induced diabetes mellitus with proliferative diabetic retinopathy with traction retinal detachment involving the macula, bilateral |
| *E093529* | Drug or chemical induced diabetes mellitus with proliferative diabetic retinopathy with traction retinal detachment involving the macula, unspecified eye |
| *E093531* | Drug or chemical induced diabetes mellitus with proliferative diabetic retinopathy with traction retinal detachment not involving the macula, right eye |
| *E093532* | Drug or chemical induced diabetes mellitus with proliferative diabetic retinopathy with traction retinal detachment not involving the macula, left eye |
| *E093533* | Drug or chemical induced diabetes mellitus with proliferative diabetic retinopathy with traction retinal detachment not involving the macula, bilateral |
| *E093539* | Drug or chemical induced diabetes mellitus with proliferative diabetic retinopathy with traction retinal detachment not involving the macula, unspecified eye |
| *E093541* | Drug or chemical induced diabetes mellitus with proliferative diabetic retinopathy with combined traction retinal detachment and rhegmatogenous retinal detachment, right eye |
| *E093542* | Drug or chemical induced diabetes mellitus with proliferative diabetic retinopathy with combined traction retinal detachment and rhegmatogenous retinal detachment, left eye |
| *E093543* | Drug or chemical induced diabetes mellitus with proliferative diabetic retinopathy with combined traction retinal detachment and rhegmatogenous retinal detachment, bilateral |
| *E093549* | Drug or chemical induced diabetes mellitus with proliferative diabetic retinopathy with combined traction retinal detachment and rhegmatogenous retinal detachment, unspecified eye |
| *E093551* | Drug or chemical induced diabetes mellitus with stable proliferative diabetic retinopathy, right eye |
| *E093552* | Drug or chemical induced diabetes mellitus with stable proliferative diabetic retinopathy, left eye |
| *E093553* | Drug or chemical induced diabetes mellitus with stable proliferative diabetic retinopathy, bilateral |
| *E093559* | Drug or chemical induced diabetes mellitus with stable proliferative diabetic retinopathy, unspecified eye |
| *E09359* | Drug or chemical induced diabetes mellitus with proliferative diabetic retinopathy without macular edema |
| *E093591* | Drug or chemical induced diabetes mellitus with proliferative diabetic retinopathy without macular edema, right eye |
| *E093592* | Drug or chemical induced diabetes mellitus with proliferative diabetic retinopathy without macular edema, left eye |
| *E093593* | Drug or chemical induced diabetes mellitus with proliferative diabetic retinopathy without macular edema, bilateral |
| *E093599* | Drug or chemical induced diabetes mellitus with proliferative diabetic retinopathy without macular edema, unspecified eye |
| *E0936* | Drug or chemical induced diabetes mellitus with diabetic cataract |
| *E0937X1* | Drug or chemical induced diabetes mellitus with diabetic macular edema, resolved following treatment, right eye |
| *E0937X2* | Drug or chemical induced diabetes mellitus with diabetic macular edema, resolved following treatment, left eye |
| *E0937X3* | Drug or chemical induced diabetes mellitus with diabetic macular edema, resolved following treatment, bilateral |
| *E0937X9* | Drug or chemical induced diabetes mellitus with diabetic macular edema, resolved following treatment, unspecified eye |
| *E0939* | Drug or chemical induced diabetes mellitus with other diabetic ophthalmic complication |
| *E0940* | Drug or chemical induced diabetes mellitus with neurological complications with diabetic neuropathy, unspecified |
| *E0941* | Drug or chemical induced diabetes mellitus with neurological complications with diabetic mononeuropathy |
| *E0942* | Drug or chemical induced diabetes mellitus with neurological complications with diabetic polyneuropathy |
| *E0943* | Drug or chemical induced diabetes mellitus with neurological complications with diabetic autonomic (poly)neuropathy |
| *E0944* | Drug or chemical induced diabetes mellitus with neurological complications with diabetic amyotrophy |
| *E0949* | Drug or chemical induced diabetes mellitus with neurological complications with other diabetic neurological complication |
| *E0951* | Drug or chemical induced diabetes mellitus with diabetic peripheral angiopathy without gangrene |
| *E0952* | Drug or chemical induced diabetes mellitus with diabetic peripheral angiopathy with gangrene |
| *E0959* | Drug or chemical induced diabetes mellitus with other circulatory complications |
| *E09610* | Drug or chemical induced diabetes mellitus with diabetic neuropathic arthropathy |
| *E09618* | Drug or chemical induced diabetes mellitus with other diabetic arthropathy |
| *E09620* | Drug or chemical induced diabetes mellitus with diabetic dermatitis |
| *E09621* | Drug or chemical induced diabetes mellitus with foot ulcer |
| *E09622* | Drug or chemical induced diabetes mellitus with other skin ulcer |
| *E09628* | Drug or chemical induced diabetes mellitus with other skin complications |
| *E09630* | Drug or chemical induced diabetes mellitus with periodontal disease |
| *E09638* | Drug or chemical induced diabetes mellitus with other oral complications |
| *E09641* | Drug or chemical induced diabetes mellitus with hypoglycemia with coma |
| *E09649* | Drug or chemical induced diabetes mellitus with hypoglycemia without coma |
| *E0965* | Drug or chemical induced diabetes mellitus with hyperglycemia |
| *E0969* | Drug or chemical induced diabetes mellitus with other specified complication |
| *E098* | Drug or chemical induced diabetes mellitus with unspecified complications |
| *E1010* | Type 1 diabetes mellitus with ketoacidosis without coma |
| *E1011* | Type 1 diabetes mellitus with ketoacidosis with coma |
| *E1021* | Type 1 diabetes mellitus with diabetic nephropathy |
| *E1022* | Type 1 diabetes mellitus with diabetic chronic kidney disease |
| *E1029* | Type 1 diabetes mellitus with other diabetic kidney complication |
| *E10311* | Type 1 diabetes mellitus with unspecified diabetic retinopathy with macular edema |
| *E10319* | Type 1 diabetes mellitus with unspecified diabetic retinopathy without macular edema |
| *E10321* | Type 1 diabetes mellitus with mild nonproliferative diabetic retinopathy with macular edema |
| *E103211* | Type 1 diabetes mellitus with mild nonproliferative diabetic retinopathy with macular edema, right eye |
| *E103212* | Type 1 diabetes mellitus with mild nonproliferative diabetic retinopathy with macular edema, left eye |
| *E103213* | Type 1 diabetes mellitus with mild nonproliferative diabetic retinopathy with macular edema, bilateral |
| *E103219* | Type 1 diabetes mellitus with mild nonproliferative diabetic retinopathy with macular edema, unspecified eye |
| *E10329* | Type 1 diabetes mellitus with mild nonproliferative diabetic retinopathy without macular edema |
| *E103291* | Type 1 diabetes mellitus with mild nonproliferative diabetic retinopathy without macular edema, right eye |
| *E103292* | Type 1 diabetes mellitus with mild nonproliferative diabetic retinopathy without macular edema, left eye |
| *E103293* | Type 1 diabetes mellitus with mild nonproliferative diabetic retinopathy without macular edema, bilateral |
| *E103299* | Type 1 diabetes mellitus with mild nonproliferative diabetic retinopathy without macular edema, unspecified eye |
| *E10331* | Type 1 diabetes mellitus with moderate nonproliferative diabetic retinopathy with macular edema |
| *E103311* | Type 1 diabetes mellitus with moderate nonproliferative diabetic retinopathy with macular edema, right eye |
| *E103312* | Type 1 diabetes mellitus with moderate nonproliferative diabetic retinopathy with macular edema, left eye |
| *E103313* | Type 1 diabetes mellitus with moderate nonproliferative diabetic retinopathy with macular edema, bilateral |
| *E103319* | Type 1 diabetes mellitus with moderate nonproliferative diabetic retinopathy with macular edema, unspecified eye |
| *E10339* | Type 1 diabetes mellitus with moderate nonproliferative diabetic retinopathy without macular edema |
| *E103391* | Type 1 diabetes mellitus with moderate nonproliferative diabetic retinopathy without macular edema, right eye |
| *E103392* | Type 1 diabetes mellitus with moderate nonproliferative diabetic retinopathy without macular edema, left eye |
| *E103393* | Type 1 diabetes mellitus with moderate nonproliferative diabetic retinopathy without macular edema, bilateral |
| *E103399* | Type 1 diabetes mellitus with moderate nonproliferative diabetic retinopathy without macular edema, unspecified eye |
| *E10341* | Type 1 diabetes mellitus with severe nonproliferative diabetic retinopathy with macular edema |
| *E103411* | Type 1 diabetes mellitus with severe nonproliferative diabetic retinopathy with macular edema, right eye |
| *E103412* | Type 1 diabetes mellitus with severe nonproliferative diabetic retinopathy with macular edema, left eye |
| *E103413* | Type 1 diabetes mellitus with severe nonproliferative diabetic retinopathy with macular edema, bilateral |
| *E103419* | Type 1 diabetes mellitus with severe nonproliferative diabetic retinopathy with macular edema, unspecified eye |
| *E10349* | Type 1 diabetes mellitus with severe nonproliferative diabetic retinopathy without macular edema |
| *E103491* | Type 1 diabetes mellitus with severe nonproliferative diabetic retinopathy without macular edema, right eye |
| *E103492* | Type 1 diabetes mellitus with severe nonproliferative diabetic retinopathy without macular edema, left eye |
| *E103493* | Type 1 diabetes mellitus with severe nonproliferative diabetic retinopathy without macular edema, bilateral |
| *E103499* | Type 1 diabetes mellitus with severe nonproliferative diabetic retinopathy without macular edema, unspecified eye |
| *E10351* | Type 1 diabetes mellitus with proliferative diabetic retinopathy with macular edema |
| *E103511* | Type 1 diabetes mellitus with proliferative diabetic retinopathy with macular edema, right eye |
| *E103512* | Type 1 diabetes mellitus with proliferative diabetic retinopathy with macular edema, left eye |
| *E103513* | Type 1 diabetes mellitus with proliferative diabetic retinopathy with macular edema, bilateral |
| *E103519* | Type 1 diabetes mellitus with proliferative diabetic retinopathy with macular edema, unspecified eye |
| *E103521* | Type 1 diabetes mellitus with proliferative diabetic retinopathy with traction retinal detachment involving the macula, right eye |
| *E103522* | Type 1 diabetes mellitus with proliferative diabetic retinopathy with traction retinal detachment involving the macula, left eye |
| *E103523* | Type 1 diabetes mellitus with proliferative diabetic retinopathy with traction retinal detachment involving the macula, bilateral |
| *E103529* | Type 1 diabetes mellitus with proliferative diabetic retinopathy with traction retinal detachment involving the macula, unspecified eye |
| *E103531* | Type 1 diabetes mellitus with proliferative diabetic retinopathy with traction retinal detachment not involving the macula, right eye |
| *E103532* | Type 1 diabetes mellitus with proliferative diabetic retinopathy with traction retinal detachment not involving the macula, left eye |
| *E103533* | Type 1 diabetes mellitus with proliferative diabetic retinopathy with traction retinal detachment not involving the macula, bilateral |
| *E103539* | Type 1 diabetes mellitus with proliferative diabetic retinopathy with traction retinal detachment not involving the macula, unspecified eye |
| *E103541* | Type 1 diabetes mellitus with proliferative diabetic retinopathy with combined traction retinal detachment and rhegmatogenous retinal detachment, right eye |
| *E103542* | Type 1 diabetes mellitus with proliferative diabetic retinopathy with combined traction retinal detachment and rhegmatogenous retinal detachment, left eye |
| *E103543* | Type 1 diabetes mellitus with proliferative diabetic retinopathy with combined traction retinal detachment and rhegmatogenous retinal detachment, bilateral |
| *E103549* | Type 1 diabetes mellitus with proliferative diabetic retinopathy with combined traction retinal detachment and rhegmatogenous retinal detachment, unspecified eye |
| *E103551* | Type 1 diabetes mellitus with stable proliferative diabetic retinopathy, right eye |
| *E103552* | Type 1 diabetes mellitus with stable proliferative diabetic retinopathy, left eye |
| *E103553* | Type 1 diabetes mellitus with stable proliferative diabetic retinopathy, bilateral |
| *E103559* | Type 1 diabetes mellitus with stable proliferative diabetic retinopathy, unspecified eye |
| *E10359* | Type 1 diabetes mellitus with proliferative diabetic retinopathy without macular edema |
| *E103591* | Type 1 diabetes mellitus with proliferative diabetic retinopathy without macular edema, right eye |
| *E103592* | Type 1 diabetes mellitus with proliferative diabetic retinopathy without macular edema, left eye |
| *E103593* | Type 1 diabetes mellitus with proliferative diabetic retinopathy without macular edema, bilateral |
| *E103599* | Type 1 diabetes mellitus with proliferative diabetic retinopathy without macular edema, unspecified eye |
| *E1036* | Type 1 diabetes mellitus with diabetic cataract |
| *E1037X1* | Type 1 diabetes mellitus with diabetic macular edema, resolved following treatment, right eye |
| *E1037X2* | Type 1 diabetes mellitus with diabetic macular edema, resolved following treatment, left eye |
| *E1037X3* | Type 1 diabetes mellitus with diabetic macular edema, resolved following treatment, bilateral |
| *E1037X9* | Type 1 diabetes mellitus with diabetic macular edema, resolved following treatment, unspecified eye |
| *E1039* | Type 1 diabetes mellitus with other diabetic ophthalmic complication |
| *E1040* | Type 1 diabetes mellitus with diabetic neuropathy, unspecified |
| *E1041* | Type 1 diabetes mellitus with diabetic mononeuropathy |
| *E1042* | Type 1 diabetes mellitus with diabetic polyneuropathy |
| *E1043* | Type 1 diabetes mellitus with diabetic autonomic (poly)neuropathy |
| *E1044* | Type 1 diabetes mellitus with diabetic amyotrophy |
| *E1049* | Type 1 diabetes mellitus with other diabetic neurological complication |
| *E1051* | Type 1 diabetes mellitus with diabetic peripheral angiopathy without gangrene |
| *E1052* | Type 1 diabetes mellitus with diabetic peripheral angiopathy with gangrene |
| *E1059* | Type 1 diabetes mellitus with other circulatory complications |
| *E10610* | Type 1 diabetes mellitus with diabetic neuropathic arthropathy |
| *E10618* | Type 1 diabetes mellitus with other diabetic arthropathy |
| *E10620* | Type 1 diabetes mellitus with diabetic dermatitis |
| *E10621* | Type 1 diabetes mellitus with foot ulcer |
| *E10622* | Type 1 diabetes mellitus with other skin ulcer |
| *E10628* | Type 1 diabetes mellitus with other skin complications |
| *E10630* | Type 1 diabetes mellitus with periodontal disease |
| *E10638* | Type 1 diabetes mellitus with other oral complications |
| *E10641* | Type 1 diabetes mellitus with hypoglycemia with coma |
| *E10649* | Type 1 diabetes mellitus with hypoglycemia without coma |
| *E1065* | Type 1 diabetes mellitus with hyperglycemia |
| *E1069* | Type 1 diabetes mellitus with other specified complication |
| *E108* | Type 1 diabetes mellitus with unspecified complications |
| *E1100* | Type 2 diabetes mellitus with hyperosmolarity without nonketotic hyperglycemic-hyperosmolar coma (NKHHC) |
| *E1101* | Type 2 diabetes mellitus with hyperosmolarity with coma |
| *E1110* | Type 2 diabetes mellitus with ketoacidosis without coma |
| *E1111* | Type 2 diabetes mellitus with ketoacidosis with coma |
| *E1121* | Type 2 diabetes mellitus with diabetic nephropathy |
| *E1122* | Type 2 diabetes mellitus with diabetic chronic kidney disease |
| *E1129* | Type 2 diabetes mellitus with other diabetic kidney complication |
| *E11311* | Type 2 diabetes mellitus with unspecified diabetic retinopathy with macular edema |
| *E11319* | Type 2 diabetes mellitus with unspecified diabetic retinopathy without macular edema |
| *E11321* | Type 2 diabetes mellitus with mild nonproliferative diabetic retinopathy with macular edema |
| *E113211* | Type 2 diabetes mellitus with mild nonproliferative diabetic retinopathy with macular edema, right eye |
| *E113212* | Type 2 diabetes mellitus with mild nonproliferative diabetic retinopathy with macular edema, left eye |
| *E113213* | Type 2 diabetes mellitus with mild nonproliferative diabetic retinopathy with macular edema, bilateral |
| *E113219* | Type 2 diabetes mellitus with mild nonproliferative diabetic retinopathy with macular edema, unspecified eye |
| *E11329* | Type 2 diabetes mellitus with mild nonproliferative diabetic retinopathy without macular edema |
| *E113291* | Type 2 diabetes mellitus with mild nonproliferative diabetic retinopathy without macular edema, right eye |
| *E113292* | Type 2 diabetes mellitus with mild nonproliferative diabetic retinopathy without macular edema, left eye |
| *E113293* | Type 2 diabetes mellitus with mild nonproliferative diabetic retinopathy without macular edema, bilateral |
| *E113299* | Type 2 diabetes mellitus with mild nonproliferative diabetic retinopathy without macular edema, unspecified eye |
| *E11331* | Type 2 diabetes mellitus with moderate nonproliferative diabetic retinopathy with macular edema |
| *E113311* | Type 2 diabetes mellitus with moderate nonproliferative diabetic retinopathy with macular edema, right eye |
| *E113312* | Type 2 diabetes mellitus with moderate nonproliferative diabetic retinopathy with macular edema, left eye |
| *E113313* | Type 2 diabetes mellitus with moderate nonproliferative diabetic retinopathy with macular edema, bilateral |
| *E113319* | Type 2 diabetes mellitus with moderate nonproliferative diabetic retinopathy with macular edema, unspecified eye |
| *E11339* | Type 2 diabetes mellitus with moderate nonproliferative diabetic retinopathy without macular edema |
| *E113391* | Type 2 diabetes mellitus with moderate nonproliferative diabetic retinopathy without macular edema, right eye |
| *E113392* | Type 2 diabetes mellitus with moderate nonproliferative diabetic retinopathy without macular edema, left eye |
| *E113393* | Type 2 diabetes mellitus with moderate nonproliferative diabetic retinopathy without macular edema, bilateral |
| *E113399* | Type 2 diabetes mellitus with moderate nonproliferative diabetic retinopathy without macular edema, unspecified eye |
| *E11341* | Type 2 diabetes mellitus with severe nonproliferative diabetic retinopathy with macular edema |
| *E113411* | Type 2 diabetes mellitus with severe nonproliferative diabetic retinopathy with macular edema, right eye |
| *E113412* | Type 2 diabetes mellitus with severe nonproliferative diabetic retinopathy with macular edema, left eye |
| *E113413* | Type 2 diabetes mellitus with severe nonproliferative diabetic retinopathy with macular edema, bilateral |
| *E113419* | Type 2 diabetes mellitus with severe nonproliferative diabetic retinopathy with macular edema, unspecified eye |
| *E11349* | Type 2 diabetes mellitus with severe nonproliferative diabetic retinopathy without macular edema |
| *E113491* | Type 2 diabetes mellitus with severe nonproliferative diabetic retinopathy without macular edema, right eye |
| *E113492* | Type 2 diabetes mellitus with severe nonproliferative diabetic retinopathy without macular edema, left eye |
| *E113493* | Type 2 diabetes mellitus with severe nonproliferative diabetic retinopathy without macular edema, bilateral |
| *E113499* | Type 2 diabetes mellitus with severe nonproliferative diabetic retinopathy without macular edema, unspecified eye |
| *E11351* | Type 2 diabetes mellitus with proliferative diabetic retinopathy with macular edema |
| *E113511* | Type 2 diabetes mellitus with proliferative diabetic retinopathy with macular edema, right eye |
| *E113512* | Type 2 diabetes mellitus with proliferative diabetic retinopathy with macular edema, left eye |
| *E113513* | Type 2 diabetes mellitus with proliferative diabetic retinopathy with macular edema, bilateral |
| *E113519* | Type 2 diabetes mellitus with proliferative diabetic retinopathy with macular edema, unspecified eye |
| *E113521* | Type 2 diabetes mellitus with proliferative diabetic retinopathy with traction retinal detachment involving the macula, right eye |
| *E113522* | Type 2 diabetes mellitus with proliferative diabetic retinopathy with traction retinal detachment involving the macula, left eye |
| *E113523* | Type 2 diabetes mellitus with proliferative diabetic retinopathy with traction retinal detachment involving the macula, bilateral |
| *E113529* | Type 2 diabetes mellitus with proliferative diabetic retinopathy with traction retinal detachment involving the macula, unspecified eye |
| *E113531* | Type 2 diabetes mellitus with proliferative diabetic retinopathy with traction retinal detachment not involving the macula, right eye |
| *E113532* | Type 2 diabetes mellitus with proliferative diabetic retinopathy with traction retinal detachment not involving the macula, left eye |
| *E113533* | Type 2 diabetes mellitus with proliferative diabetic retinopathy with traction retinal detachment not involving the macula, bilateral |
| *E113539* | Type 2 diabetes mellitus with proliferative diabetic retinopathy with traction retinal detachment not involving the macula, unspecified eye |
| *E113541* | Type 2 diabetes mellitus with proliferative diabetic retinopathy with combined traction retinal detachment and rhegmatogenous retinal detachment, right eye |
| *E113542* | Type 2 diabetes mellitus with proliferative diabetic retinopathy with combined traction retinal detachment and rhegmatogenous retinal detachment, left eye |
| *E113543* | Type 2 diabetes mellitus with proliferative diabetic retinopathy with combined traction retinal detachment and rhegmatogenous retinal detachment, bilateral |
| *E113549* | Type 2 diabetes mellitus with proliferative diabetic retinopathy with combined traction retinal detachment and rhegmatogenous retinal detachment, unspecified eye |
| *E113551* | Type 2 diabetes mellitus with stable proliferative diabetic retinopathy, right eye |
| *E113552* | Type 2 diabetes mellitus with stable proliferative diabetic retinopathy, left eye |
| *E113553* | Type 2 diabetes mellitus with stable proliferative diabetic retinopathy, bilateral |
| *E113559* | Type 2 diabetes mellitus with stable proliferative diabetic retinopathy, unspecified eye |
| *E11359* | Type 2 diabetes mellitus with proliferative diabetic retinopathy without macular edema |
| *E113591* | Type 2 diabetes mellitus with proliferative diabetic retinopathy without macular edema, right eye |
| *E113592* | Type 2 diabetes mellitus with proliferative diabetic retinopathy without macular edema, left eye |
| *E113593* | Type 2 diabetes mellitus with proliferative diabetic retinopathy without macular edema, bilateral |
| *E113599* | Type 2 diabetes mellitus with proliferative diabetic retinopathy without macular edema, unspecified eye |
| *E1136* | Type 2 diabetes mellitus with diabetic cataract |
| *E1137X1* | Type 2 diabetes mellitus with diabetic macular edema, resolved following treatment, right eye |
| *E1137X2* | Type 2 diabetes mellitus with diabetic macular edema, resolved following treatment, left eye |
| *E1137X3* | Type 2 diabetes mellitus with diabetic macular edema, resolved following treatment, bilateral |
| *E1137X9* | Type 2 diabetes mellitus with diabetic macular edema, resolved following treatment, unspecified eye |
| *E1139* | Type 2 diabetes mellitus with other diabetic ophthalmic complication |
| *E1140* | Type 2 diabetes mellitus with diabetic neuropathy, unspecified |
| *E1141* | Type 2 diabetes mellitus with diabetic mononeuropathy |
| *E1142* | Type 2 diabetes mellitus with diabetic polyneuropathy |
| *E1143* | Type 2 diabetes mellitus with diabetic autonomic (poly)neuropathy |
| *E1144* | Type 2 diabetes mellitus with diabetic amyotrophy |
| *E1149* | Type 2 diabetes mellitus with other diabetic neurological complication |
| *E1151* | Type 2 diabetes mellitus with diabetic peripheral angiopathy without gangrene |
| *E1152* | Type 2 diabetes mellitus with diabetic peripheral angiopathy with gangrene |
| *E1159* | Type 2 diabetes mellitus with other circulatory complications |
| *E11610* | Type 2 diabetes mellitus with diabetic neuropathic arthropathy |
| *E11618* | Type 2 diabetes mellitus with other diabetic arthropathy |
| *E11620* | Type 2 diabetes mellitus with diabetic dermatitis |
| *E11621* | Type 2 diabetes mellitus with foot ulcer |
| *E11622* | Type 2 diabetes mellitus with other skin ulcer |
| *E11628* | Type 2 diabetes mellitus with other skin complications |
| *E11630* | Type 2 diabetes mellitus with periodontal disease |
| *E11638* | Type 2 diabetes mellitus with other oral complications |
| *E11641* | Type 2 diabetes mellitus with hypoglycemia with coma |
| *E11649* | Type 2 diabetes mellitus with hypoglycemia without coma |
| *E1165* | Type 2 diabetes mellitus with hyperglycemia |
| *E1169* | Type 2 diabetes mellitus with other specified complication |
| *E118* | Type 2 diabetes mellitus with unspecified complications |
| *E1300* | Other specified diabetes mellitus with hyperosmolarity without nonketotic hyperglycemic-hyperosmolar coma (NKHHC) |
| *E1301* | Other specified diabetes mellitus with hyperosmolarity with coma |
| *E1310* | Other specified diabetes mellitus with ketoacidosis without coma |
| *E1311* | Other specified diabetes mellitus with ketoacidosis with coma |
| *E1321* | Other specified diabetes mellitus with diabetic nephropathy |
| *E1322* | Other specified diabetes mellitus with diabetic chronic kidney disease |
| *E1329* | Other specified diabetes mellitus with other diabetic kidney complication |
| *E13311* | Other specified diabetes mellitus with unspecified diabetic retinopathy with macular edema |
| *E13319* | Other specified diabetes mellitus with unspecified diabetic retinopathy without macular edema |
| *E13321* | Other specified diabetes mellitus with mild nonproliferative diabetic retinopathy with macular edema |
| *E133211* | Other specified diabetes mellitus with mild nonproliferative diabetic retinopathy with macular edema, right eye |
| *E133212* | Other specified diabetes mellitus with mild nonproliferative diabetic retinopathy with macular edema, left eye |
| *E133213* | Other specified diabetes mellitus with mild nonproliferative diabetic retinopathy with macular edema, bilateral |
| *E133219* | Other specified diabetes mellitus with mild nonproliferative diabetic retinopathy with macular edema, unspecified eye |
| *E13329* | Other specified diabetes mellitus with mild nonproliferative diabetic retinopathy without macular edema |
| *E133291* | Other specified diabetes mellitus with mild nonproliferative diabetic retinopathy without macular edema, right eye |
| *E133292* | Other specified diabetes mellitus with mild nonproliferative diabetic retinopathy without macular edema, left eye |
| *E133293* | Other specified diabetes mellitus with mild nonproliferative diabetic retinopathy without macular edema, bilateral |
| *E133299* | Other specified diabetes mellitus with mild nonproliferative diabetic retinopathy without macular edema, unspecified eye |
| *E13331* | Other specified diabetes mellitus with moderate nonproliferative diabetic retinopathy with macular edema |
| *E133311* | Other specified diabetes mellitus with moderate nonproliferative diabetic retinopathy with macular edema, right eye |
| *E133312* | Other specified diabetes mellitus with moderate nonproliferative diabetic retinopathy with macular edema, left eye |
| *E133313* | Other specified diabetes mellitus with moderate nonproliferative diabetic retinopathy with macular edema, bilateral |
| *E133319* | Other specified diabetes mellitus with moderate nonproliferative diabetic retinopathy with macular edema, unspecified eye |
| *E13339* | Other specified diabetes mellitus with moderate nonproliferative diabetic retinopathy without macular edema |
| *E133391* | Other specified diabetes mellitus with moderate nonproliferative diabetic retinopathy without macular edema, right eye |
| *E133392* | Other specified diabetes mellitus with moderate nonproliferative diabetic retinopathy without macular edema, left eye |
| *E133393* | Other specified diabetes mellitus with moderate nonproliferative diabetic retinopathy without macular edema, bilateral |
| *E133399* | Other specified diabetes mellitus with moderate nonproliferative diabetic retinopathy without macular edema, unspecified eye |
| *E13341* | Other specified diabetes mellitus with severe nonproliferative diabetic retinopathy with macular edema |
| *E133411* | Other specified diabetes mellitus with severe nonproliferative diabetic retinopathy with macular edema, right eye |
| *E133412* | Other specified diabetes mellitus with severe nonproliferative diabetic retinopathy with macular edema, left eye |
| *E133413* | Other specified diabetes mellitus with severe nonproliferative diabetic retinopathy with macular edema, bilateral |
| *E133419* | Other specified diabetes mellitus with severe nonproliferative diabetic retinopathy with macular edema, unspecified eye |
| *E13349* | Other specified diabetes mellitus with severe nonproliferative diabetic retinopathy without macular edema |
| *E133491* | Other specified diabetes mellitus with severe nonproliferative diabetic retinopathy without macular edema, right eye |
| *E133492* | Other specified diabetes mellitus with severe nonproliferative diabetic retinopathy without macular edema, left eye |
| *E133493* | Other specified diabetes mellitus with severe nonproliferative diabetic retinopathy without macular edema, bilateral |
| *E133499* | Other specified diabetes mellitus with severe nonproliferative diabetic retinopathy without macular edema, unspecified eye |
| *E13351* | Other specified diabetes mellitus with proliferative diabetic retinopathy with macular edema |
| *E133511* | Other specified diabetes mellitus with proliferative diabetic retinopathy with macular edema, right eye |
| *E133512* | Other specified diabetes mellitus with proliferative diabetic retinopathy with macular edema, left eye |
| *E133513* | Other specified diabetes mellitus with proliferative diabetic retinopathy with macular edema, bilateral |
| *E133519* | Other specified diabetes mellitus with proliferative diabetic retinopathy with macular edema, unspecified eye |
| *E133521* | Other specified diabetes mellitus with proliferative diabetic retinopathy with traction retinal detachment involving the macula, right eye |
| *E133522* | Other specified diabetes mellitus with proliferative diabetic retinopathy with traction retinal detachment involving the macula, left eye |
| *E133523* | Other specified diabetes mellitus with proliferative diabetic retinopathy with traction retinal detachment involving the macula, bilateral |
| *E133529* | Other specified diabetes mellitus with proliferative diabetic retinopathy with traction retinal detachment involving the macula, unspecified eye |
| *E133531* | Other specified diabetes mellitus with proliferative diabetic retinopathy with traction retinal detachment not involving the macula, right eye |
| *E133532* | Other specified diabetes mellitus with proliferative diabetic retinopathy with traction retinal detachment not involving the macula, left eye |
| *E133533* | Other specified diabetes mellitus with proliferative diabetic retinopathy with traction retinal detachment not involving the macula, bilateral |
| *E133539* | Other specified diabetes mellitus with proliferative diabetic retinopathy with traction retinal detachment not involving the macula, unspecified eye |
| *E133541* | Other specified diabetes mellitus with proliferative diabetic retinopathy with combined traction retinal detachment and rhegmatogenous retinal detachment, right eye |
| *E133542* | Other specified diabetes mellitus with proliferative diabetic retinopathy with combined traction retinal detachment and rhegmatogenous retinal detachment, left eye |
| *E133543* | Other specified diabetes mellitus with proliferative diabetic retinopathy with combined traction retinal detachment and rhegmatogenous retinal detachment, bilateral |
| *E133549* | Other specified diabetes mellitus with proliferative diabetic retinopathy with combined traction retinal detachment and rhegmatogenous retinal detachment, unspecified eye |
| *E133551* | Other specified diabetes mellitus with stable proliferative diabetic retinopathy, right eye |
| *E133552* | Other specified diabetes mellitus with stable proliferative diabetic retinopathy, left eye |
| *E133553* | Other specified diabetes mellitus with stable proliferative diabetic retinopathy, bilateral |
| *E133559* | Other specified diabetes mellitus with stable proliferative diabetic retinopathy, unspecified eye |
| *E13359* | Other specified diabetes mellitus with proliferative diabetic retinopathy without macular edema |
| *E133591* | Other specified diabetes mellitus with proliferative diabetic retinopathy without macular edema, right eye |
| *E133592* | Other specified diabetes mellitus with proliferative diabetic retinopathy without macular edema, left eye |
| *E133593* | Other specified diabetes mellitus with proliferative diabetic retinopathy without macular edema, bilateral |
| *E133599* | Other specified diabetes mellitus with proliferative diabetic retinopathy without macular edema, unspecified eye |
| *E1336* | Other specified diabetes mellitus with diabetic cataract |
| *E1337X1* | Type 2 diabetes mellitus with diabetic macular edema, resolved following treatment, right eye |
| *E1337X2* | Type 2 diabetes mellitus with diabetic macular edema, resolved following treatment, left eye |
| *E1337X3* | Type 2 diabetes mellitus with diabetic macular edema, resolved following treatment, bilateral |
| *E1337X9* | Type 2 diabetes mellitus with diabetic macular edema, resolved following treatment, unspecified eye |
| *E1339* | Other specified diabetes mellitus with other diabetic ophthalmic complication |
| *E1340* | Other specified diabetes mellitus with diabetic neuropathy, unspecified |
| *E1341* | Other specified diabetes mellitus with diabetic mononeuropathy |
| *E1342* | Other specified diabetes mellitus with diabetic polyneuropathy |
| *E1343* | Other specified diabetes mellitus with diabetic autonomic (poly)neuropathy |
| *E1344* | Other specified diabetes mellitus with diabetic amyotrophy |
| *E1349* | Other specified diabetes mellitus with other diabetic neurological complication |
| *E1351* | Other specified diabetes mellitus with diabetic peripheral angiopathy without gangrene |
| *E1352* | Other specified diabetes mellitus with diabetic peripheral angiopathy with gangrene |
| *E1359* | Other specified diabetes mellitus with other circulatory complications |
| *E13610* | Other specified diabetes mellitus with diabetic neuropathic arthropathy |
| *E13618* | Other specified diabetes mellitus with other diabetic arthropathy |
| *E13620* | Other specified diabetes mellitus with diabetic dermatitis |
| *E13621* | Other specified diabetes mellitus with foot ulcer |
| *E13622* | Other specified diabetes mellitus with other skin ulcer |
| *E13628* | Other specified diabetes mellitus with other skin complications |
| *E13630* | Other specified diabetes mellitus with periodontal disease |
| *E13638* | Other specified diabetes mellitus with other oral complications |
| *E13641* | Other specified diabetes mellitus with hypoglycemia with coma |
| *E13649* | Other specified diabetes mellitus with hypoglycemia without coma |
| *E1365* | Other specified diabetes mellitus with hyperglycemia |
| *E1369* | Other specified diabetes mellitus with other specified complication |
| *E138* | Other specified diabetes mellitus with unspecified complications |
| *G3289* | Other specified degenerative disorders of nervous system in diseases classified elsewhere |
| *E089* | Diabetes mellitus due to underlying condition without complications |
| *E099* | Drug or chemical induced diabetes mellitus without complications |
| *E109* | Type 1 diabetes mellitus without complications |
| *E119* | Type 2 diabetes mellitus without complications |
| *E139* | Other specified diabetes mellitus without complications |
| *R7301* | Impaired fasting glucose |
| *R7302* | Impaired glucose tolerance (oral) |
| R7303 | Prediabetes |
| R7309 | Other abnormal glucose |
| R739 | Hyperglycemia, unspecified |
| R81 | Glycosuria |
| R824 | Acetonuria |
| Z4681 | Encounter for fitting and adjustment of insulin pump |
| Z9641 | Presence of insulin pump (external) (internal) |

##### Hypertension ICD 10

|  |  |
| --- | --- |
| I10 | *Essential (primary) hypertension* |
| I110 | Hypertensive heart disease with heart failure |
| I119 | Hypertensive heart disease without heart failure |
| I120 | Hypertensive chronic kidney disease with stage 5 chronic kidney disease or end stage renal disease |
| I129 | Hypertensive chronic kidney disease with stage 1 through stage 4 chronic kidney disease, or unspecified chronic kidney disease |
| I130 | Hypertensive heart and chronic kidney disease with heart failure and stage 1 through stage 4 chronic kidney disease, or unspecified chronic kidney disease |
| I1310 | Hypertensive heart and chronic kidney disease without heart failure, with stage 1 through stage 4 chronic kidney disease, or unspecified chronic kidney disease |
| I1311 | Hypertensive heart and chronic kidney disease without heart failure, with stage 5 chronic kidney disease, or end stage renal disease |
| I132 | Hypertensive heart and chronic kidney disease with heart failure and with stage 5 chronic kidney disease, or end stage renal disease |
| I150 | Renovascular hypertension |
| I151 | Hypertension secondary to other renal disorders |
| I152 | Hypertension secondary to endocrine disorders |
| I158 | Other secondary hypertension |
| I159 | Secondary hypertension, unspecified |
| I160 | Hypertensive urgency |
| I161 | Hypertensive emergency |
| I169 | Hypertensive crisis, unspecified |
| I674 | Hypertensive encephalopathy |
| N262 | Page kidney |

##### Mood disorders. ICD 10

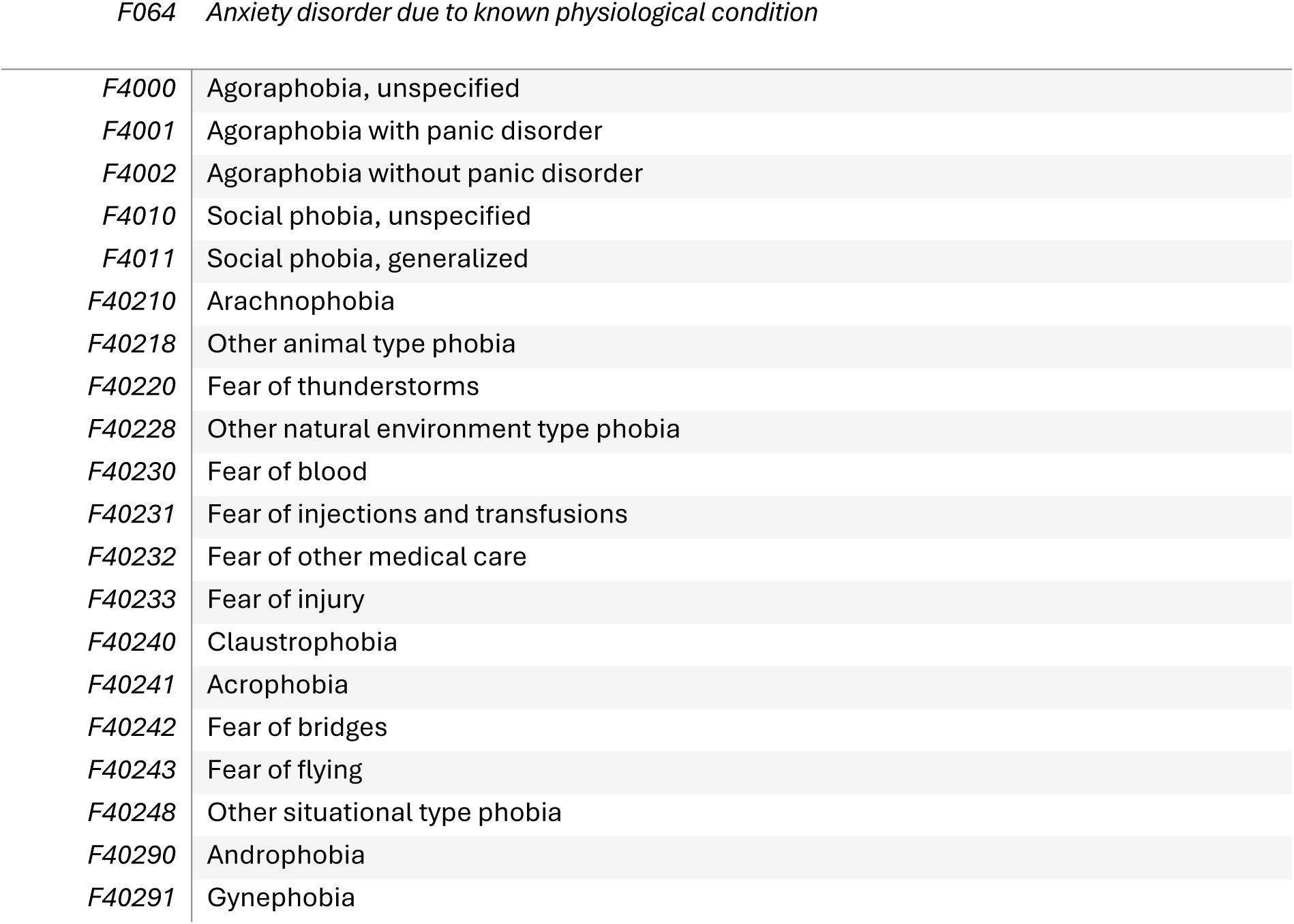
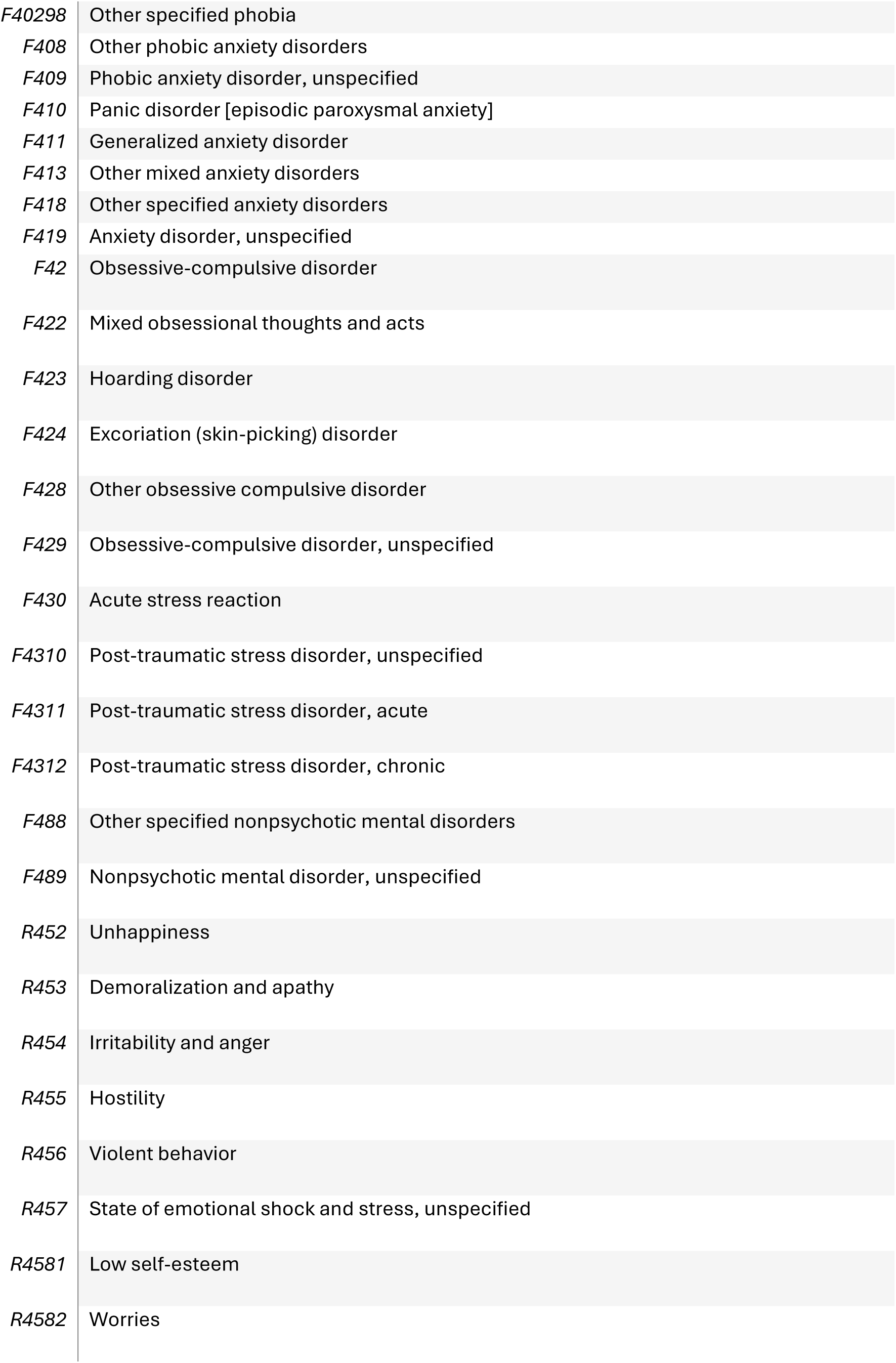
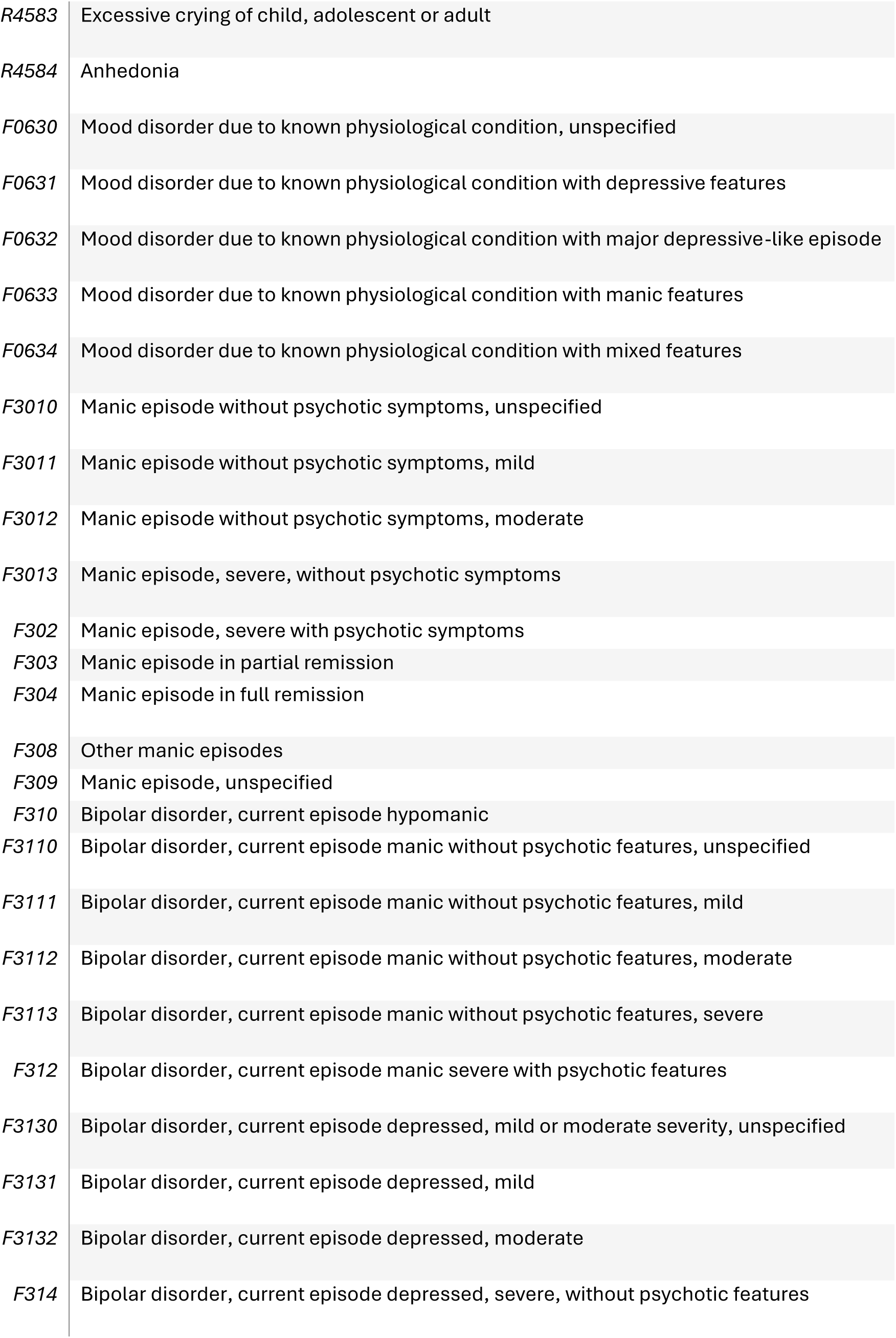
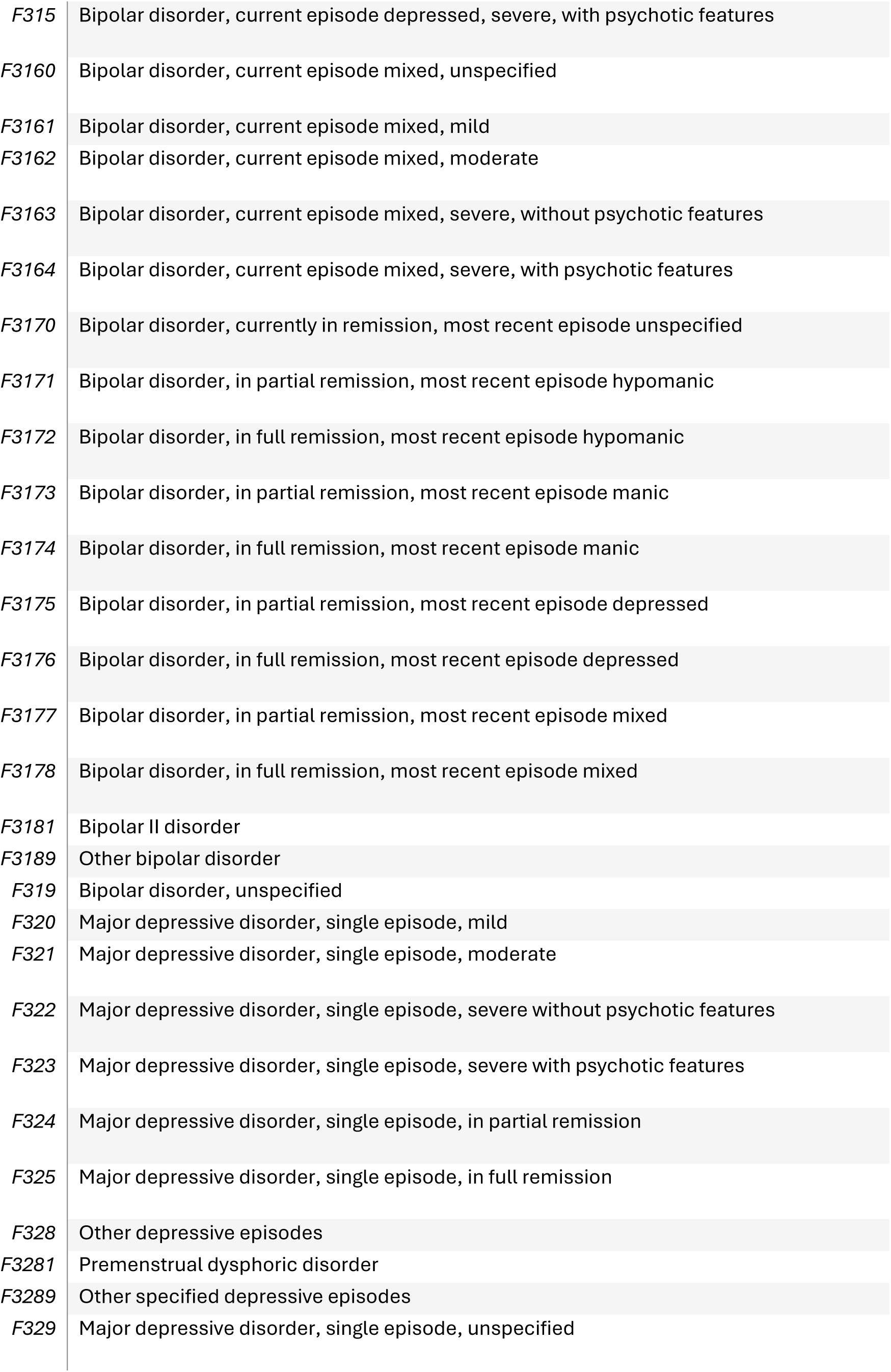
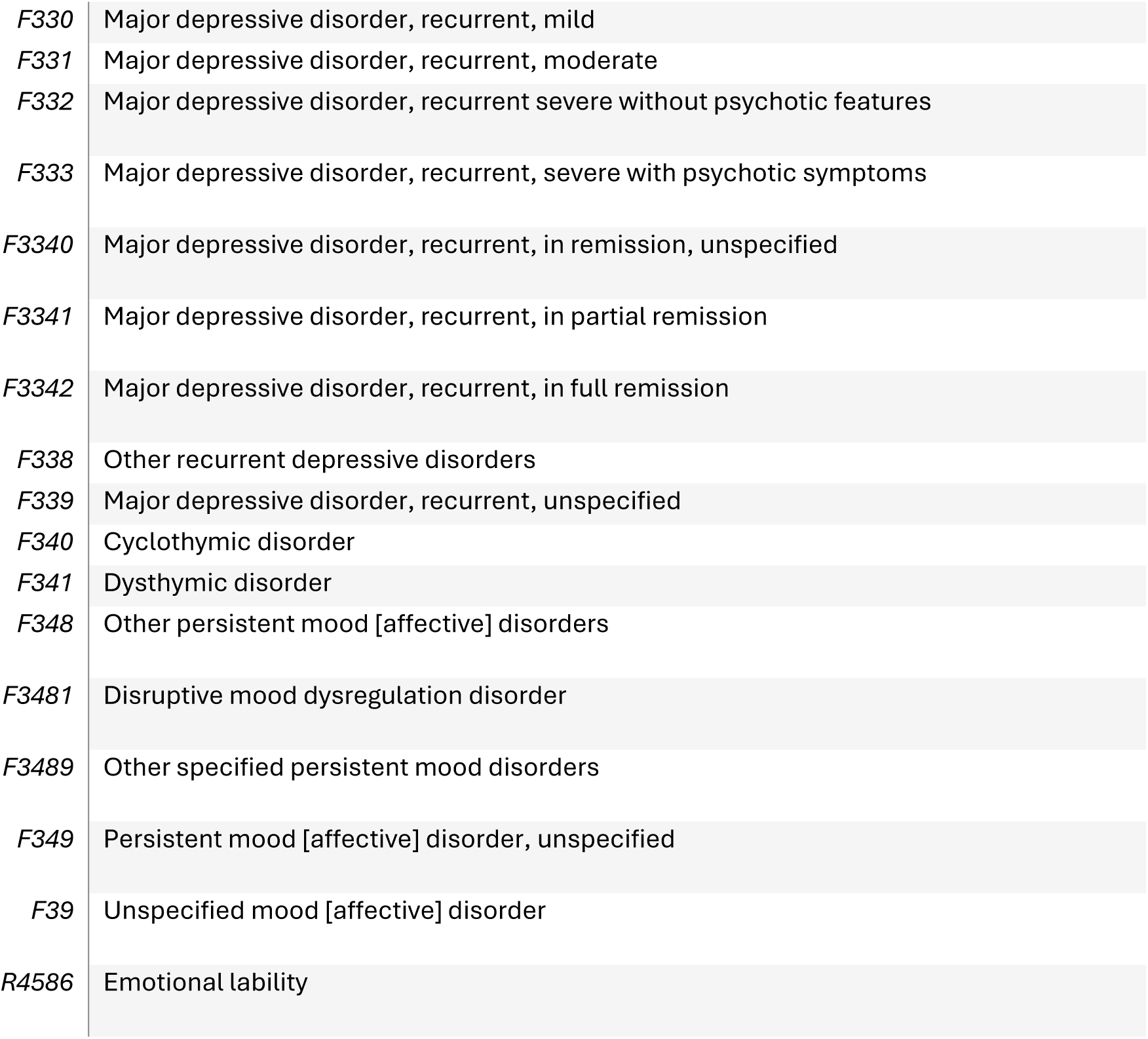

##### Asthna ICD 10

Asthma ICD 10
|  |  |
| --- | --- |
| *J4520* | *Mild intermittent asthma, uncomplicated* |
| *J4521* | Mild intermittent asthma with (acute) exacerbation |
| *J4522* | Mild intermittent asthma with status asthmaticus |
| *J4530* | Mild persistent asthma, uncomplicated |
| *J4531* | Mild persistent asthma with (acute) exacerbation |
| *J4532* | Mild persistent asthma with status asthmaticus |
| *J4540* | Moderate persistent asthma, uncomplicated |
| *J4541* | Moderate persistent asthma with (acute) exacerbation |
| *J4542* | Moderate persistent asthma with status asthmaticus |
| *J4550* | Severe persistent asthma, uncomplicated |
| *J4551* | Severe persistent asthma with (acute) exacerbation |
| *J4552* | Severe persistent asthma with status asthmaticus |
| *J45901* | Unspecified asthma with (acute) exacerbation |
| *J45902* | Unspecified asthma with status asthmaticus |
| *J45909* | Unspecified asthma, uncomplicated |
| *J45990* | Exercise induced bronchospasm |
| *J45991* | Cough variant asthma |
| *J45998* | Other asthma |

##### Coronary heart disease. ICD 10

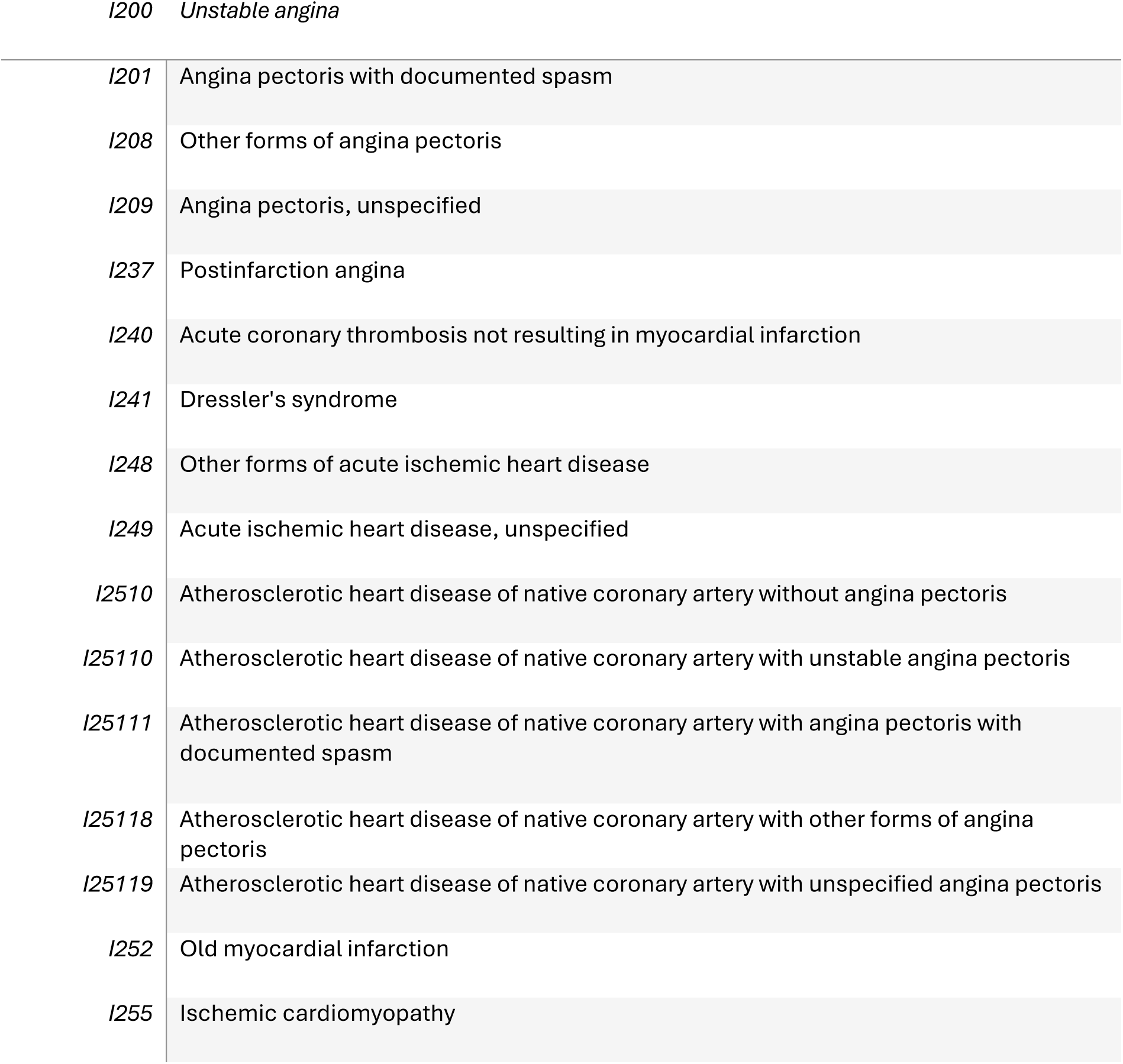
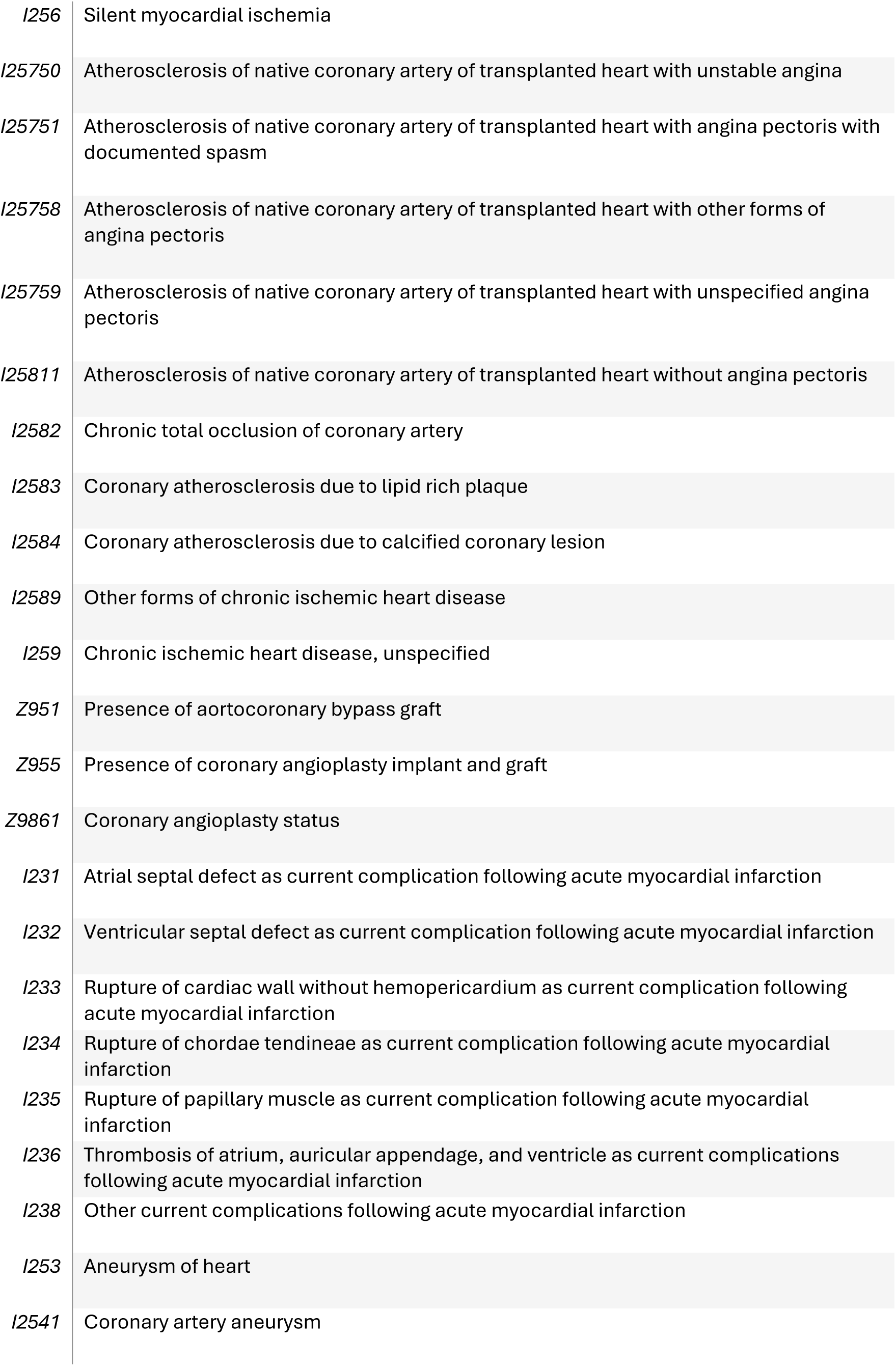
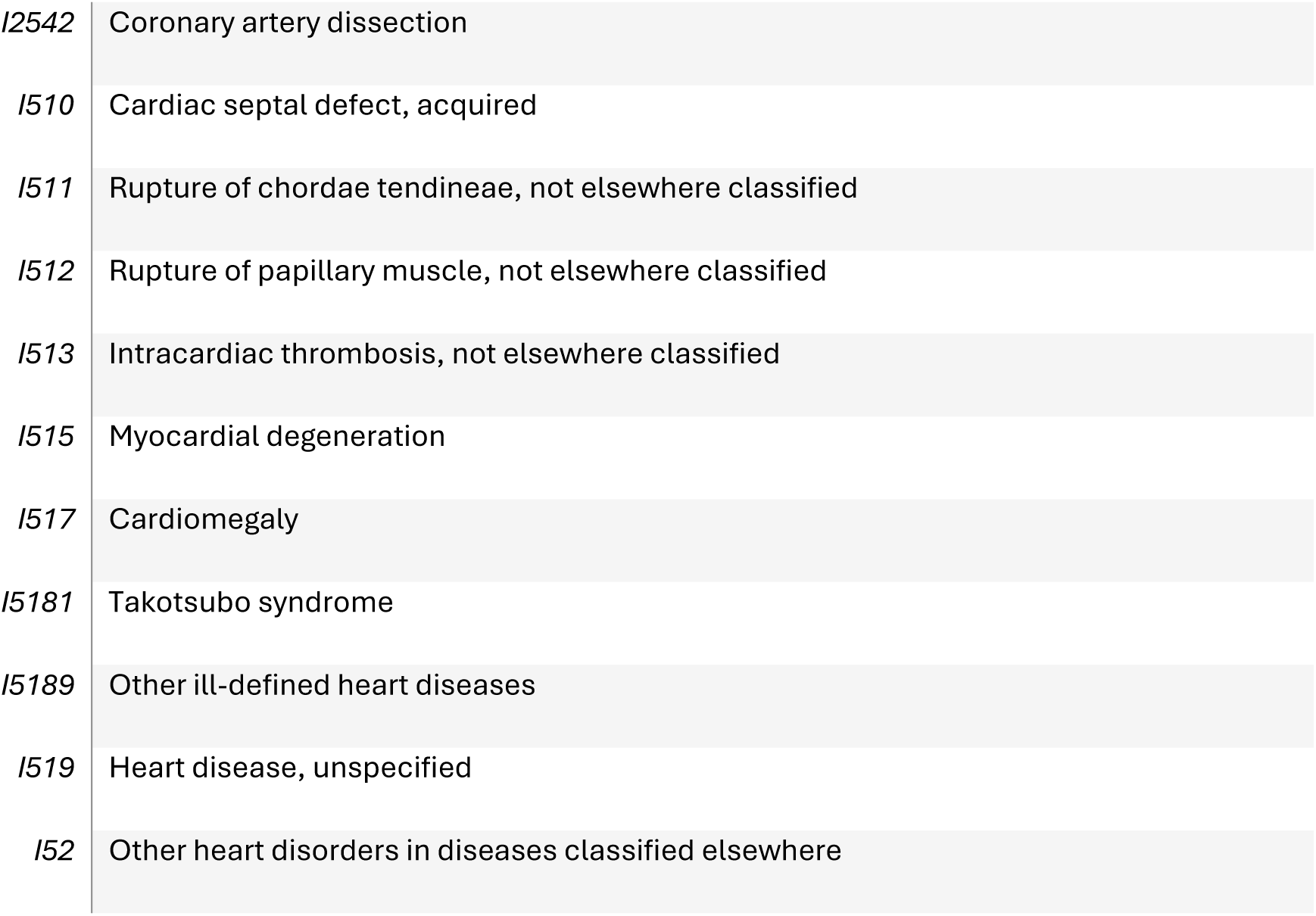

#### ICD 9

##### Diabetes ICD 9

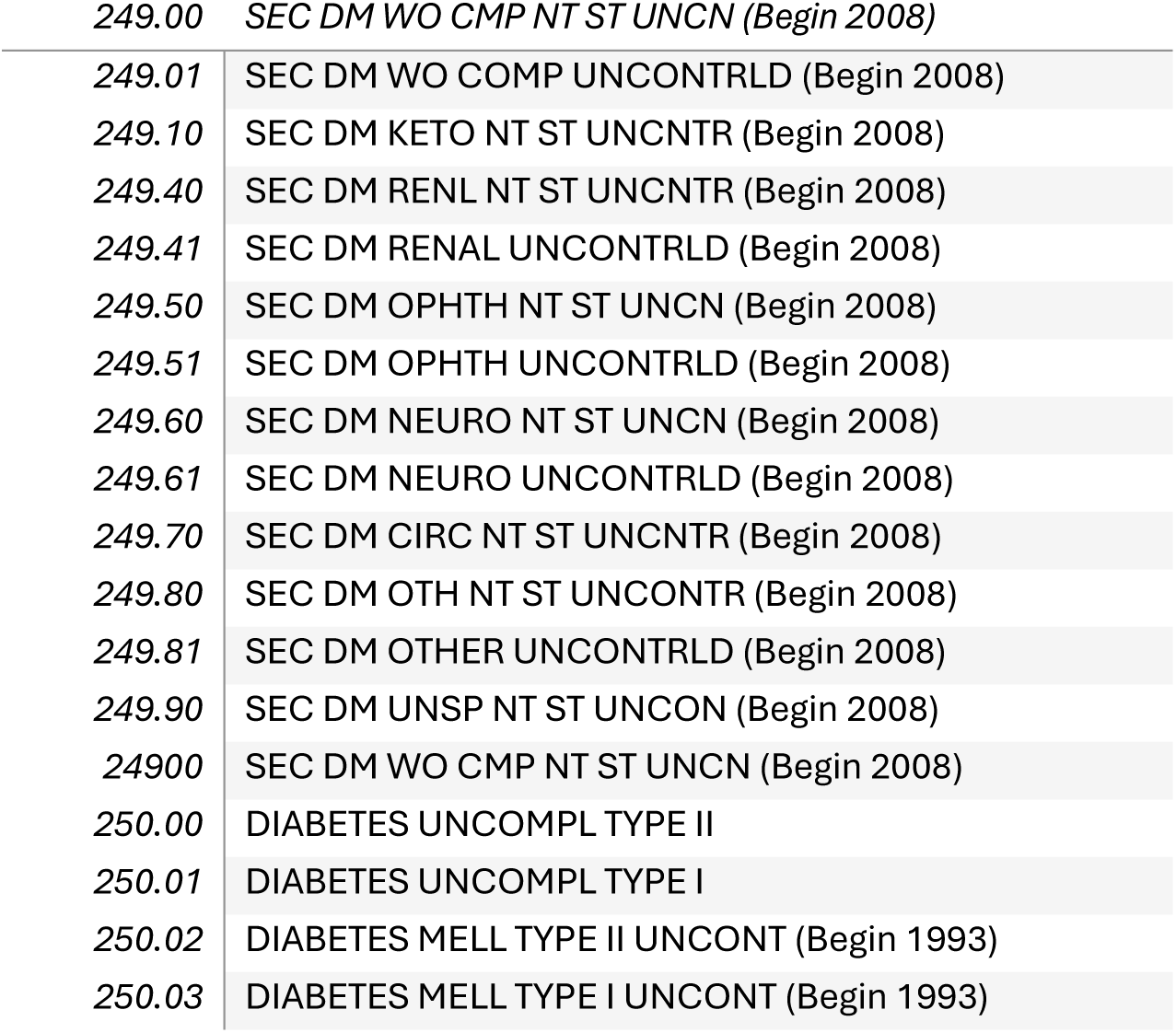
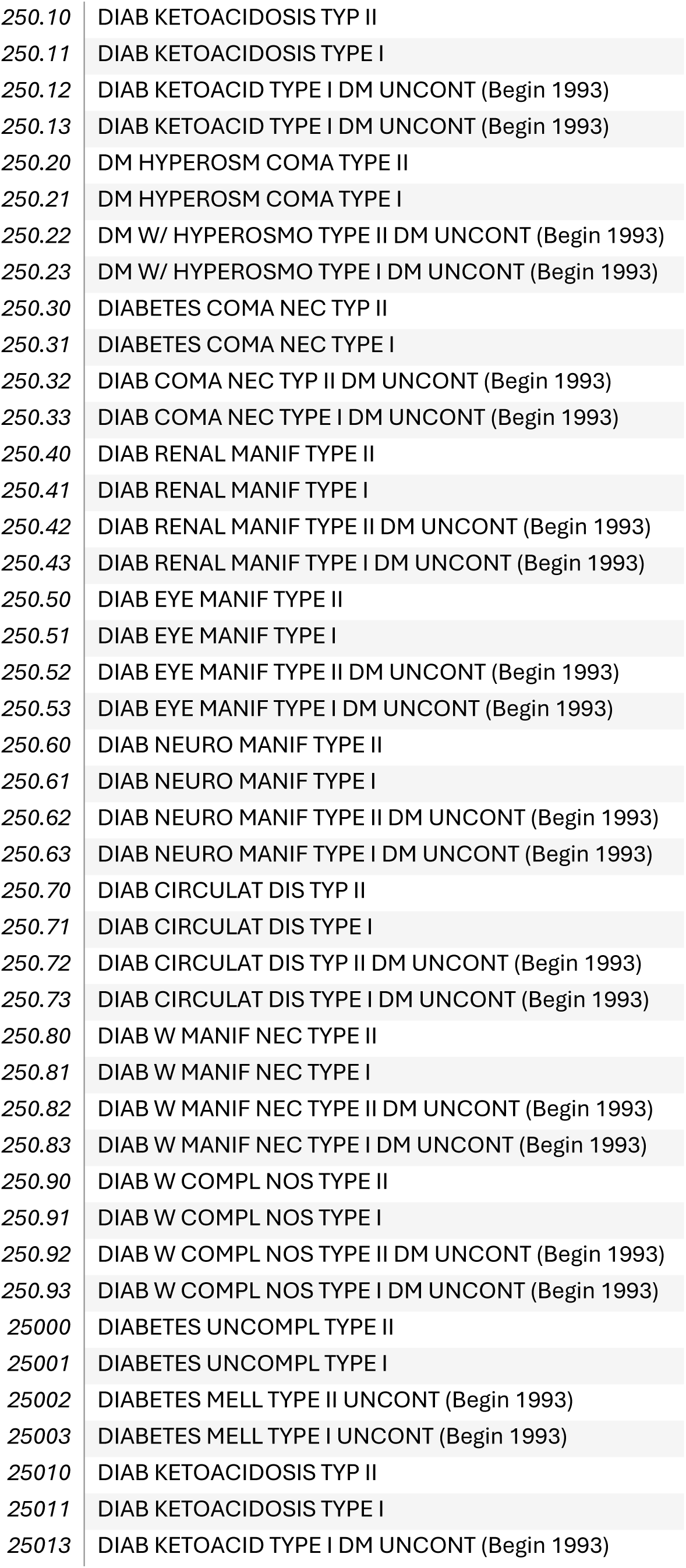
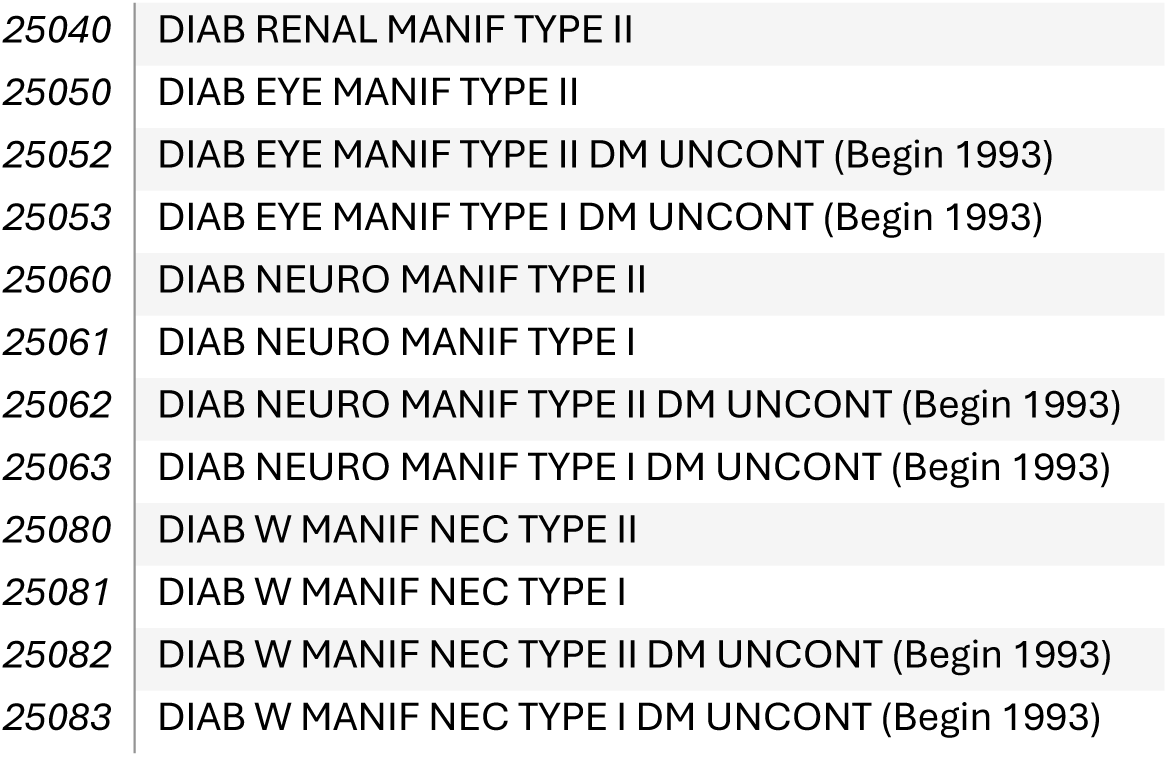

##### Hypertension ICD 9

|  |  |
| --- | --- |
| 403.1 | *BENIGN HYPERT RENAL DIS (Begin 1980 End 1989)* |
| 403.10 | BEN HYP REN W/O REN FAIL (Begin 1989) |
| 403.11 | BEN HYP RENAL W REN FAIL (Begin 1989) |
| 403.90 | HYP REN NOS W/O REN FAIL (Begin 1989) |
| 403.91 | HYP RENAL NOS W REN FAIL (Begin 1989) |
| 404.00 | MAL HY HT/REN W/O CHF/RF (Begin 1989) |
| 404.01 | MAL HYPER HRT/REN W CHF (Begin 1989) |
| 404.02 | MAL HY HT/REN W REN FAIL (Begin 1989) |
| 404.03 | MAL HYP HRT/REN W CHF & RF (Begin 1989) |
| 404.10 | BEN HY HT/REN W/O CHF/RF (Begin 1989) |
| 404.12 | BEN HY HT/REN W REN FAIL (Begin 1989) |
| 404.13 | BEN HYP HRT/REN W CHF & RF (Begin 1989) |
| 404.90 | HY HT/REN NOS W/O CHF/RF (Begin 1989) |
| 404.91 | HYPER HRT/REN NOS W CHF (Begin 1989) |
| 404.92 | HY HT/REN NOS W REN FAIL (Begin 1989) |
| 404.93 | HYP HT/REN NOS W CHF & RF (Begin 1989) |
| 405.01 | MAL RENOVASC HYPERTENS |
| 405.09 | MAL SECOND HYPERTEN NEC |
| 405.11 | BENIGN RENOVASC HYPERTEN |
| 405.19 | BENIGN SECOND HYPERT NEC |
| 405.91 | RENOVASC HYPERTENSION |
| 405.99 | SECOND HYPERTENSION NEC |
| 437.2 | HYPERTENS ENCEPHALOPATHY |

##### CHD ICD 9

|  |  |
| --- | --- |
| 411.0 | POST MI SYNDROME |
| 411.1 | INTERMED CORONARY SYND |
| 411.81 | CORONARY OCCLSN W/O MI (Begin 1989) |
| 411.89 | AC ISCHEMIC HRT DIS NEC (Begin 1989) |
| 412 | OLD MYOCARDIAL INFARCT |
| 413.0 | ANGINA DECUBITUS |
| 413.1 | PRINZMETAL ANGINA |
| 413.9 | ANGINA PECTORIS NEC/NOS |
| 414.00 | CORONARY ATHERO NOS (Begin 1994) |
| 414.01 | CORONARY ATHERO NATIVE VESSEL (Begin 1994) |
| 414.06 | CORONARY ATHERO CRNRY ARTERY OF TRANS (Begin 2002) |
| 414.10 | ANEURYSM- HEART (WALL) |
| 414.11 | CORONARY VESSEL ANEURYSM |
| 414.12 | DISSECTION OF CORONARY ARTERY (Begin 2002) |
| 414.19 | ANEURYSM OF HEART NEC |
| 414.2 | CHR TOT OCCLUS COR ARTRY (Begin 2007) |
| 414.3 | COR ATH D/T LPD RCH PLAQ (Begin 2008) |
| 414.8 | CHR ISCHEMIC HRT DIS NEC |
| 414.9 | CHR ISCHEMIC HRT DIS NOS |
| 429.1 | MYOCARDIAL DEGENERATION |
| 429.2 | ASCVD |
| 429.3 | CARDIOMEGALY |
| 429.5 | CHORDAE TENDINAE RUPTURE |
| 429.6 | PAPILLARY MUSCLE RUPTURE |
| 429.71 | ACQ CARDIAC SEPTL DEFECT (Begin 1989) |
| 429.79 | OTHER SEQUELAE OF MI NEC (Begin 1989) |
| 429.81 | PAPILLARY MUSCLE DIS NEC |
| 429.82 | HYPERKINETIC HEART DIS |
| 429.83 | TAKOTSUBO SYNDROME (Begin 2006) |
| 429.89 | ILL-DEFINED HRT DIS NEC |
| 429.9 | HEART DISEASE NOS |
| V45.81 | AORTOCORONARY BYPASS |
| V45.82 | PTCA STATUS (Begin 1994) |

##### Asthna ICD 9

Asthma ICD 9
|  |  |
| --- | --- |
| 493.00 | EXT ASTHMA W/O STAT ASTH |
| 493.01 | EXT ASTHMA W STATUS ASTH |
| 493.02 | EXT ASTHMA W/ ACUTE EXACERBATION (Begin 2000) |
| 493.10 | INT ASTHMA W/O STAT ASTH |
| 493.11 | INT ASTHMA W STATUS ASTH |
| 493.12 | INT ASTHMA W/ ACUTE EXACERBATION (Begin 2000) |
| 493.20 | CH OB ASTH W/O STAT ASTH (Begin 1989) |
| 493.21 | CH OB ASTHMA W STAT ASTH (Begin 1989) |
| 493.22 | CH OB ASTHMA W/ACUTE EXACERBATION (Begin 2000) |
| 493.81 | EXERCISE INDUCED BRONCHOSPASM (Begin 2003) |
| 493.82 | COUGH VARIANT ASTHMA (Begin 2003) |
| 493.90 | ASTHMA W/O STATUS ASTHM |
| 493.91 | ASTHMA W/ STATUS ASTHMAT |
| 493.92 | ASTHMA W/ ACUTE EXACERBATION (Begin 2000) |

##### Mood disorders ICD 9

|  |  |
| --- | --- |
| 296.11 | RECUR MANIC DIS-MILD |
| 296.12 | RECUR MANIC DIS-MOD |
| 296.13 | RECUR MANIC DIS-SEVERE |
| 296.14 | RECUR MANIC-SEV W PSYCHO |
| 296.16 | RECUR MANIC-FULL REMISS |
| 296.20 | DEPRESS PSYCHOSIS-UNSPEC |
| 296.21 | DEPRESS PSYCHOSIS-MILD |
| 296.22 | DEPRESSIVE PSYCHOSIS-MOD |
| 296.23 | DEPRESS PSYCHOSIS-SEVERE |
| 296.24 | DEPR PSYCHOS-SEV W PSYCH |
| 296.25 | DEPR PSYCHOS-PART REMISS |
| 296.26 | DEPR PSYCHOS-FULL REMISS |
| 296.30 | RECURR DEPR PSYCHOS-UNSP |
| 296.31 | RECURR DEPR PSYCHOS-MILD |
| 296.32 | RECURR DEPR PSYCHOS-MOD |
| 296.33 | RECUR DEPR PSYCH-SEVERE |
| 296.34 | REC DEPR PSYCH-PSYCHOTIC |
| 296.35 | RECUR DEPR PSYC-PART REM |
| 296.36 | RECUR DEPR PSYC-FULL REM |
| 296.40 | BIPOL AFF/ MANIC-UNSPEC |
| 296.41 | BIPOLAR AFF/ MANIC-MILD |
| 296.42 | BIPOLAR AFFEC/ MANIC-MOD |
| 296.43 | BIPOL AFF/ MANIC-SEVERE |
| 296.44 | BIPOL MANIC-SEV W PSYCH |
| 296.45 | BIPOL AFF MANIC-PART REM |
| 296.46 | BIPOL AFF MANIC-FULL REM |
| 296.50 | BIPOLAR AFF/ DEPR-UNSPEC |
| 296.51 | BIPOLAR AFFEC/ DEPR-MILD |
| 296.52 | BIPOLAR AFFEC/ DEPR-MOD |
| 296.53 | BIPOL AFF/ DEPR-SEVERE |
| 296.54 | BIPOL DEPR-SEV W PSYCH |
| 296.55 | BIPOL AFF DEPR-PART REM |
| 296.56 | BIPOL AFF DEPR-FULL REM |
| 296.60 | BIPOL AFF/ MIXED-UNSPEC |
| 296.61 | BIPOLAR AFF/ MIXED-MILD |
| 296.62 | BIPOLAR AFFEC/ MIXED-MOD |
| 296.63 | BIPOL AFF/ MIXED-SEVERE |
| 296.64 | BIPOL MIXED-SEV W PSYCH |
| 296.65 | BIPOL AFF/ MIX-PART REM |
| 296.66 | BIPOL AFF/ MIX-FULL REM |
| 296.7 | BIPOLAR AFFECTIVE NOS |
| 296.80 | MANIC-DEPRESSIVE NOS |
| 296.81 | ATYPICAL MANIC DISORDER |
| 296.82 | ATYPICAL DEPRESSIVE DIS |
| 296.89 | MANIC-DEPRESSIVE NEC |
| 296.90 | AFFECTIVE PSYCHOSIS NOS |
| 296.99 | AFFECTIVE PSYCHOSES NEC |
| 29620 | DEPRESS PSYCHOSIS-UNSPEC |
| 29623 | DEPRESS PSYCHOSIS-SEVERE |
| 29630 | RECURR DEPR PSYCHOS-UNSP |
| 29632 | RECURR DEPR PSYCHOS-MOD |
| 29633 | RECUR DEPR PSYCH-SEVERE |
| 29635 | RECUR DEPR PSYC-PART REM |
| 29640 | BIPOL AFF/ MANIC-UNSPEC |
| 29650 | BIPOLAR AFF/ DEPR-UNSPEC |
| 29651 | BIPOLAR AFFEC/ DEPR-MILD |
| 29652 | BIPOLAR AFFEC/ DEPR-MOD |
| 29654 | BIPOL DEPR-SEV W PSYCH |
| 29655 | BIPOL AFF DEPR-PART REM |
| 2967 | BIPOLAR AFFECTIVE NOS |
| 29680 | MANIC-DEPRESSIVE NOS |
| 29689 | MANIC-DEPRESSIVE NEC |
| 29690 | AFFECTIVE PSYCHOSIS NOS |
| 300.00 | ANXIETY STATE NOS |
| 300.01 | PANIC DISORDER |
| 300.02 | GENERALIZED ANXIETY DIS |
| 300.09 | ANXIETY STATE NEC |
| 300.10 | HYSTERIA NOS |
| 300.20 | PHOBIA NOS |
| 300.21 | AGORAPHOBIA WITH PANIC |
| 300.22 | AGORAPHOBIA WO PANIC |
| 300.23 | SOCIAL PHOBIA |
| 300.29 | ISOLATED PHOBIAS NEC |
| 300.3 | OBSESSIVE-COMPULSIVE DIS |
| 300.4 | NEUROTIC DEPRESSION |
| 300.5 | NEURASTHENIA |
| 300.89 | NEUROTIC DISORDERS NEC |
| 300.9 | NEUROTIC DISORDER NOS |
| 308.0 | STRESS REACT/ EMOTIONAL |
| 308.1 | STRESS REACTION/ FUGUE |
| 308.2 | STRESS REACT/ PSYCHOMOT |
| 308.3 | ACUTE STRESS REACT NEC |
| 308.4 | STRESS REACT/ MIXED DIS |
| 308.9 | ACUTE STRESS REACT NOS |
| 309.81 | PROLONG POSTTRAUM STRESS |
| 311 | DEPRESSIVE DISORDER NEC |
| 313.0 | OVERANXIOUS DISORDER |
| 313.1 | MISERY & UNHAPPINESS DIS |
| 313.21 | SHYNESS DISORDER-CHILD |
| 313.22 | INTROVERTED DIS-CHILD |
| 313.3 | RELATIONSHIP PROBLEMS |
| 313.82 | IDENTITY DISORDER |
| 313.83 | ACADEMIC UNDERACHIEVMENT |

**Table 4:** Association between each SES quartile in Urban vs rural in disease prevalence.

|  |  |
| --- | --- |
| Outcome | HOUSES x Rurality<br>Interaction p value |
| Asthma | <0.001 |
| Mood Disorder | 0.004 |
| Hypertension | 0.329 |
| Diabetes | 0.666 |
| CHD | 0.368 |

**Table 5:**
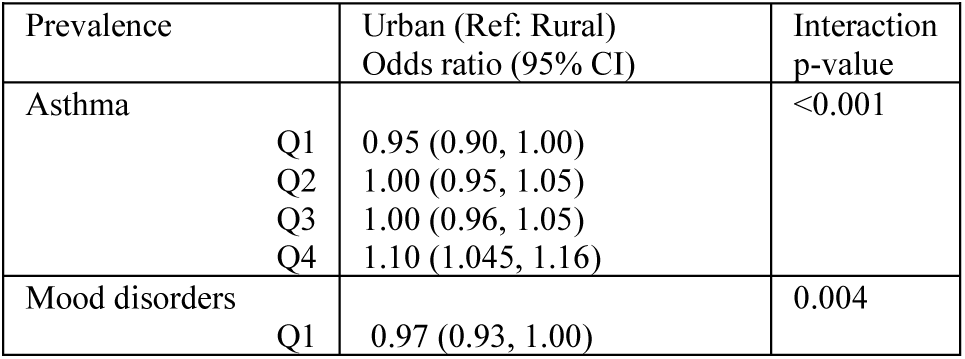
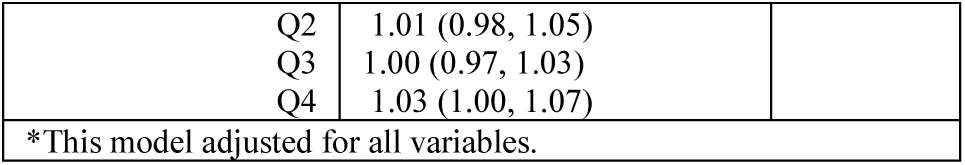
Comparison of rural and urban SES quartiles for asthma and mood disorders*.

